# A rapid review of the effectiveness of smoking cessation interventions for people with anxiety and/or depression living within the community

**DOI:** 10.1101/2024.07.23.24310849

**Authors:** Jordan Everitt, Toby Ayres, Alesha Wale, Chukwudi Okolie, Amy Fox-McNally, Helen Morgan, Hannah Shaw, Jacob Davies, Rhiannon Tudor-Edwards, Alison Cooper, Adrian Edwards, Ruth Lewis

**Affiliations:** Public Health Wales, United Kingdom; Centre for Health Economics & Medicines Evaluation (CHEME), Bangor University, United Kingdom; Health and Care Research Wales Evidence Centre, Cardiff University, United Kingdom; Health and Care Research Wales Evidence Centre, Bangor University, United Kingdom

## Abstract

The Welsh Government aims to reduce smoking prevalence from the current rate of 13% to below 5% of the population by 2030. People with mental health conditions have a higher rate of smoking prevalence and are less likely to access smoking cessation services. Evidence shows that smoking cessation in this population decreases symptoms, improves positive mood and quality of life. This rapid review aimed to identify and synthesise the evidence for the effectiveness of smoking cessation interventions in people with anxiety and/or depression living in the community.

**Results:** The literature searches were conducted in March 2024, the included study reports were published between 2008 and 2023, nine were published since 2019. Eleven primary studies from 15 reports were included in the rapid review: 10 RCTs, two of which were pilot RCTs, and one quasi-experimental pilot study. Studies were conducted in the USA (n=6), Spain (n=1), France (n=1), Netherlands (n=1), and two studies were conducted across the EU and USA.

**Research Implications and Evidence Gaps:** No UK studies were identified therefore it is unclear whether findings are generalisable to the UK. No studies applying interventions at critical touchpoints within smoking cessation or mental health services were identified. Only one study assessed the cost-effectiveness of a smoking cessation intervention. Only one study assessed a smoking cessation intervention in participants with anxiety. Most studies included in this review were judged to be of low quality. Most studies recruited participants from the general population, therefore it is unclear whether participants were engaged with mental health services. Further high-quality UK-based research is needed to better understand the effectiveness of smoking cessation interventions for people with anxiety and depression.

**Funding statement:** Public Health Wales were funded for this work by the Health and Care Research Wales Evidence Centre, itself funded by Health and Care Research Wales on behalf of Welsh Government.

**What is a Rapid Review?:** Our rapid reviews use a variation of the systematic review approach, abbreviating or omitting some components to generate the evidence to inform stakeholders promptly whilst maintaining attention to bias.

**Who is this Rapid Review for?:** The research question was suggested by Welsh Government Health Improvement.

**Background / Aim of Rapid Review:** The Welsh Government aims to reduce smoking prevalence from the current rate of 13% to below 5% of the population by 2030. People with mental health conditions have a higher rate of smoking prevalence and are less likely to access smoking cessation services. Evidence shows that smoking cessation in this population decreases symptoms, improves positive mood and quality of life. This rapid review aimed to identify and synthesise the evidence for the effectiveness of smoking cessation interventions in people with anxiety and/or depression living in the community.

**Results:** *Recency of the evidence base:*

- The literature searches were conducted in March 2024, the included study reports were published between 2008 and 2023, nine were published since 2019.

*Extent of the evidence base:*

- Eleven primary studies from 15 reports were included in the rapid review: 10 RCTs, two of which were pilot RCTs, and one quasi-experimental pilot study.
- Studies were conducted in the USA (n=6), Spain (n=1), France (n=1), Netherlands (n=1), and two studies were conducted across the EU and USA.
- Studies investigated pharmacological (Varenicline, Bupropion, nicotine replacement therapy), psychological (behavioural activation, contingency management, mood management, smoking cessation counselling), and aerobic exercise interventions. Most interventions were conducted in-person, with two studies using remote delivery via mobile applications or telephone.
- Outcomes included various measures of smoking cessation, mental health symptoms, adverse events and cost-effectiveness.

*Key findings and certainty of the evidence:*

- Overall, the evidence of effectiveness of smoking cessation interventions for those with anxiety and depression appears to be inconsistent.
- Taking into account the overall methodological quality, variability of outcome measures used and consistency of study findings, it was difficult to make direct comparison between the different studies included. Therefore, we have very low certainty across all the outcome measures identified. This means that the true effect is probably different from the estimated effect.
- There is some evidence to suggest that psychological smoking cessation interventions can increase abstinence in people with depression, however the impact on mental health outcomes appeared to be mixed.
- There is some evidence to suggest that pharmacological smoking cessation interventions can increase abstinence. However, they appeared to have no impact on mental health outcomes and no consistent impact on adverse events for people with anxiety and depression.
- Evidence shows the impact of exercise interventions had mixed findings on abstinence rates and no impact on mental health outcomes in people with depression.
- There is some evidence suggesting multicomponent pharmacological and psychological smoking cessation interventions can increase abstinence rates and reduce adverse events in people with depression. However, the impact on mental health outcomes appears mixed.
- There is limited evidence supporting the effectiveness of multicomponent exercise and psychological smoking cessation interventions but no impact on abstinence rates or mental health outcomes in people with depression.
- There is very limited evidence suggesting psychological smoking cessation interventions for people with depression may be cost-effective.

*Research Implications and Evidence Gaps:*

- No UK studies were identified therefore it is unclear whether findings are generalisable to the UK.
- No studies applying interventions at critical touchpoints within smoking cessation or mental health services were identified.
- Only one study assessed the cost-effectiveness of a smoking cessation intervention.
- Only one study assessed a smoking cessation intervention in participants with anxiety.
- Most studies included in this review were judged to be of low quality.
- Most studies recruited participants from the general population, therefore it is unclear whether participants were engaged with mental health services.
- Further high-quality UK-based research is needed to better understand the effectiveness of smoking cessation interventions for people with anxiety and depression.

*Policy and Practice Implications:*

- There is limited high quality evidence on smoking cessation interventions for those with anxiety or depression, therefore cautious consideration of findings is required if this evidence is used to inform future interventions.
- Although low quality evidence supports the use of pharmacological, psychological, and multicomponent pharmacological and psychological smoking cessation interventions to increase abstinence in people with depression.
- This review identified (limited/low quality) Varenicline as potentially effective for smoking cessation in the population group of interest. In light of recent All Wales Medicine Strategy Group recommendations to allow Cytosine, which has similar action to Varenicline, this may be of particular interest.

*Economic considerations:*

- Smoking is a considerable public health issue that incurs significant economic costs. The estimated economic cost of smoking in people with mental health disorders in the UK is £3.5 billion per annum.
- There is limited economic evidence on the impact of implementing smoking cessation interventions for individuals living with depression and/or anxiety. A summary of findings and the certainty of evidence has been assessed using an approach adapted from the GRADE evidence profile (Guyatt et al, 2011), which has been adapted for the purpose of this review.

*Disclaimer:* The views expressed in this publication are those of the authors, not necessarily Health and Care Research Wales. The Health and Care Research Wales Evidence Centre and authors of this work declare that they have no conflict of interest.

## 1. BACKGROUND

### 1.1 Who is this review for?

This Rapid Review was conducted as part of the Health and Care Research Wales Evidence Centre Work Programme. The research question was formulated by Welsh Government Health Improvement to inform the Welsh Government delivery plan for 2024-2026.

### 1.2 Background and purpose of this review

Smoking remains a significant public health concern in Wales. It is the leading cause of preventable death in Wales and costs the Welsh NHS an estimated £302 million annually (Welsh Government 2022). The Welsh Government has published a long-term plan towards a smoke-free Wales, with the aim of reducing smoking prevalence from the current rate of 13%, to below 5% of the population by 2030 (Welsh Government 2022).

People with mental health conditions, including those with anxiety and depression, have significantly higher smoking rates compared to the general population; in Wales, 35% of adults aged 16 and over who report having a long-standing mental health condition are smokers (Welsh Government, 2023a). This is over double the rate of smokers in the general population of Wales (13%) (Welsh Government 2023b). These populations are also less likely to access smoking cessation services (National Institute for Health and Care Excellence, 2021). Furthermore, the life expectancy of people with poor mental health is on average 10 to 20 years less than the general population, with smoking being the biggest cause of this difference (Public Health England, 2020).

Anxiety and depression together are the most prevalent common mental health disorders (NHS Digital, 2016). According to the most recent Adult Psychiatric Morbidity Survey, 5.9% and 3.3% of adults aged 16 years and over meet the diagnostic criteria for anxiety or depression, respectively (NHS Digital 2016). Additionally, recent data collected by the Office for National Statistics (2021), showed 16% of adults in Great Britain reported they were experiencing depressive symptoms, and 16% were also likely to be experiencing some form of anxiety.

A range of smoking cessation interventions have been shown to be effective in the general population. According to two Cochrane systematic reviews, these include behavioural interventions such as counselling and guaranteed financial incentives (Hartmann-Boyce et al, 2021), and pharmacological interventions like Varenicline, Bupropion, cytosine, nicotine replacement therapy (NRT), and nicotine e-cigarettes (Lindson et al, 2023). It is unclear if these interventions would also be effective in people with anxiety and depression. Wales’ national smoking cessation service, Help Me Quit, currently offers six weeks of behavioural support and 12 weeks of NRT to people who smoke however its effectiveness among people with anxiety and depression is largely unknown. Research suggests that people with mental health disorders are likely to have a better quality of life and feel more positive and calmer after quitting smoking (NHS, 2024). The beneficial effects of giving up smoking on anxiety and depression symptoms is said to equal that of taking anti-depressant medication (NHS, 2024). There is therefore a need to identify the most effective smoking cessation interventions for this population group.

This rapid review focuses on primary studies and aims to identify and synthesise the evidence for the effectiveness of smoking cessation interventions for people with anxiety and/or depression living in the community.

## 2. RESULTS

### 2.1 Overview of the Evidence Base

Eleven studies (15 reports) were eligible for inclusion in this rapid review. Eligibility was restricted to comparative primary studies assessing the effectiveness of smoking cessation interventions for adult smokers with anxiety or depression living in the community (see section 5.1 for full eligibility criteria). Ten of the 11 studies were randomised controlled trials (RCTs), two of which were pilot studies. One study was a quasi-experimental uncontrolled before and after pilot study. Studies were conducted in a range of countries, including USA (n=6), Spain (n=1), France (n=1), Netherlands (n=1), and two studies were conducted across multiple countries across the EU and USA. Sample sizes varied across the 15 identified reports, ranging from 17 to 6,653, and 11 reports included more than 100 participants. Reports were published between 2008 and 2023, nine since 2019.

A protocol for a UK-based study investigating the acceptability and impact of a smoking cessation intervention for mental health service users with depression, alongside routine psychological therapy was also identified during searching. This study may be particularly useful for informing smoking cessation interventions for people with anxiety and/or depression as it is UK based and is incorporated into existing psychological therapy for mental health service users. We were informed by the corresponding author that the study report is currently under consideration with the Addiction Journal. The protocol is registered on the International Standard Randomised Controlled Trial Number (ISRCTN); ISRCTN99531779 (https://doi.org/10.1186/ISRCTN99531779).

All of the studies included participants who had depression (n=11), one of which also included people with anxiety. A range of diagnostic tools were used to identify study participants with depression and anxiety at study entry. This included the Structured Clinical Interview for axis I disorders of the DSM-IV, the Beck Depression Inventory-II (BDI-II), the neuropsychiatric interview, the Centre for Epidemiological Studies Depression (CES-D) scale, the Patient Health Questionnaire (PHQ) versions 8/9, and the Hospital Anxiety and Depression Scale (HADS).

For the synthesis of results, studies have been grouped into five categories according to the type of intervention(s) being assessed: psychological (n=3), pharmacological (n=3), exercise-based (n=2), multicomponent pharmacological and psychological (n=2), and multicomponent exercise and psychological interventions (n=1). Greater detail of the specific interventions being investigated is shown in Table 1, which also outlines the current treatment used in Wales. All intervention components only received by participants in the experimental group are highlighted in green (an Asterix is also given for readers with difficulty distinguishing colours). In most studies (n=8), the interventions being assessed were an addition to existing smoking cessation treatment which was also received by participants in the control group. The smoking cessation treatment components received by all participants (intervention and control groups) are highlighted in yellow in Table 1 (or X without an Asterisk).

**Table 1.**
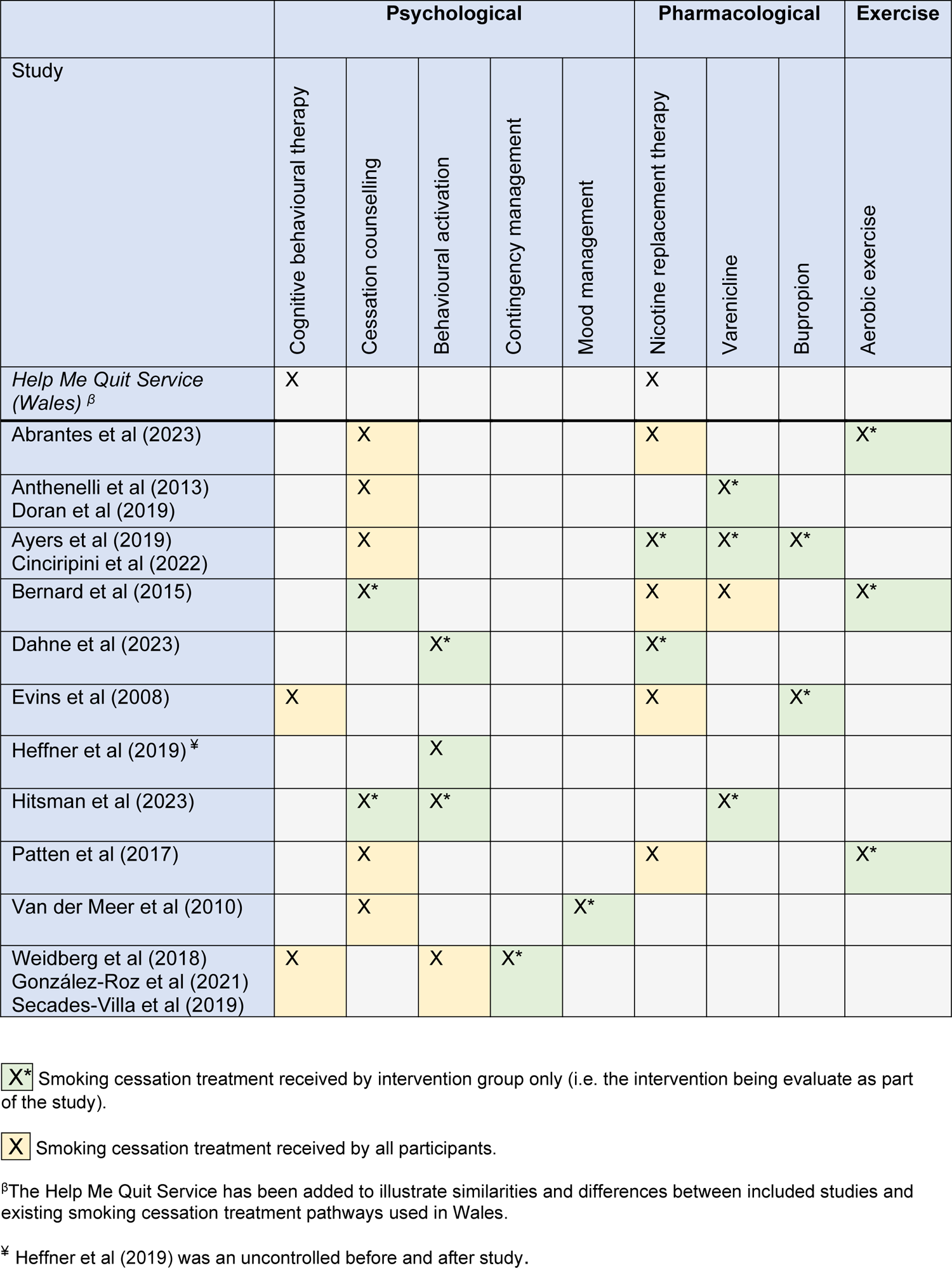
Intervention components and smoking cessation treatments received by participants in the included studies and the current treatment used in Wales.

|  | Psychological |  |  |  |  | Pharmacological |  |  | Exercise |
| --- | --- | --- | --- | --- | --- | --- | --- | --- | --- |
| Study | Cognitive behavioural therapy | Cessation counselling | Behavioural activation | Contingency management | Mood management | Nicotine replacement therapy | Varenicline | Bupropion | Aerobic exercise |
| *Help Me Quit Service (Wales)*<sup>β</sup> | X |  |  |  |  | X |  |  |  |
| Abrantes et al (2023) |  | X |  |  |  | X |  |  | X* |
| Anthenelli et al (2013)<br>Doran et al (2019) |  | X |  |  |  |  | X* |  |  |
| Ayers et al (2019)<br>Cinciripini et al (2022) |  | X |  |  |  | X* | X* | X* |  |
| Bernard et al (2015) |  | X* |  |  |  | X | X |  | X* |
| Dahne et al (2023) |  |  | X* |  |  | X* |  |  |  |
| Evins et al (2008) | X |  |  |  |  | X |  | X* |  |
| Heffner et al (2019)<sup>‡</sup> |  |  | X |  |  |  |  |  |  |
| Hitsman et al (2023) |  | X* | X* |  |  |  | X* |  |  |
| Patten et al (2017) |  | X |  |  |  | X |  |  | X* |
| Van der Meer et al (2010) |  | X |  |  | X* |  |  |  |  |
| Weidberg et al (2018)<br>González-Roz et al (2021)<br>Secades-Villa et al (2019) | X |  | X | X* |  |  |  |  |  |

**X\*** Smoking cessation treatment received by intervention group only (i.e. the intervention being evaluate as part of the study).
**X** Smoking cessation treatment received by all participants.
<sup>β</sup>The Help Me Quit Service has been added to illustrate similarities and differences between included studies and existing smoking cessation treatment pathways used in Wales.
<sup>‡</sup> Heffner et al (2019) was an uncontrolled before and after study.

Interventions were delivered at clinical trials centres or outpatient clinics (n=3), research clinics (n=2), a hospital (n=1), and remotely via a mobile phone application, text messaging programme, or telephone (n=2). Three studies did not state the intervention setting.

Intervention durations ranged from six weeks to three months. Details of the interventions can be seen in Table 2; a summary of the findings can be found in Table 3, and a more detailed summary of the included studies can be seen in section 6.2 (Table 5).

**Table 2.**
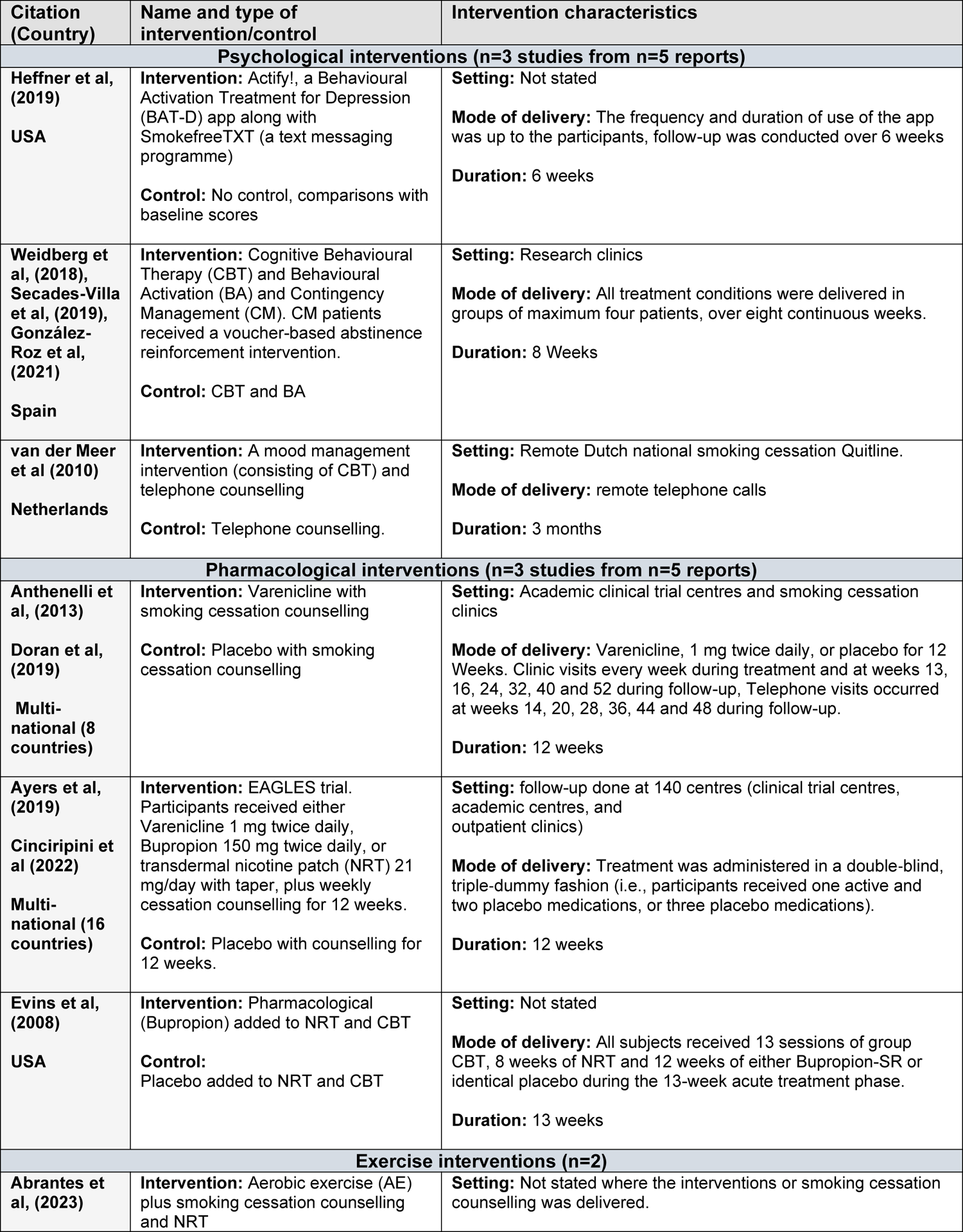
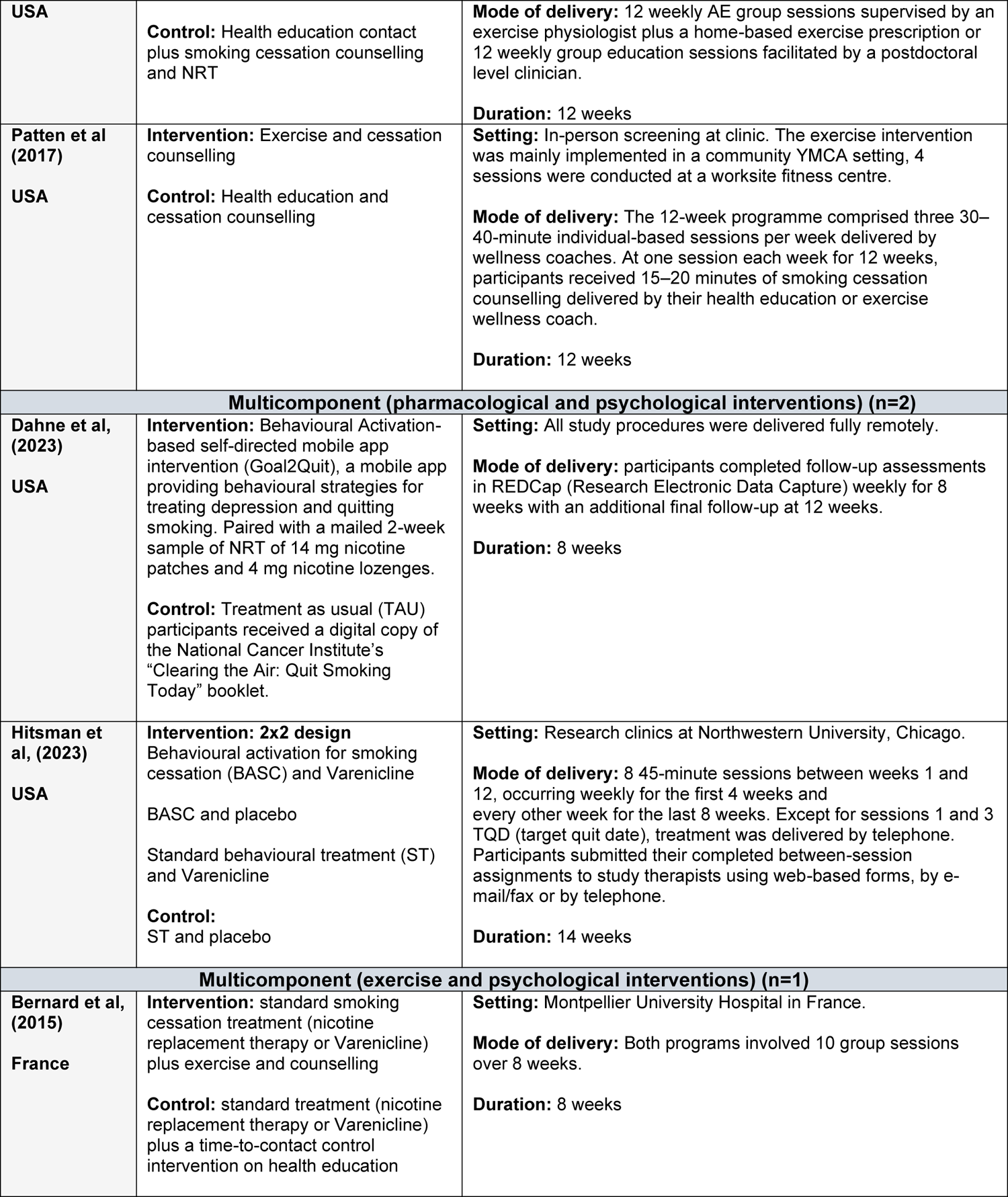
Summary of interventions.

**Table 3.**
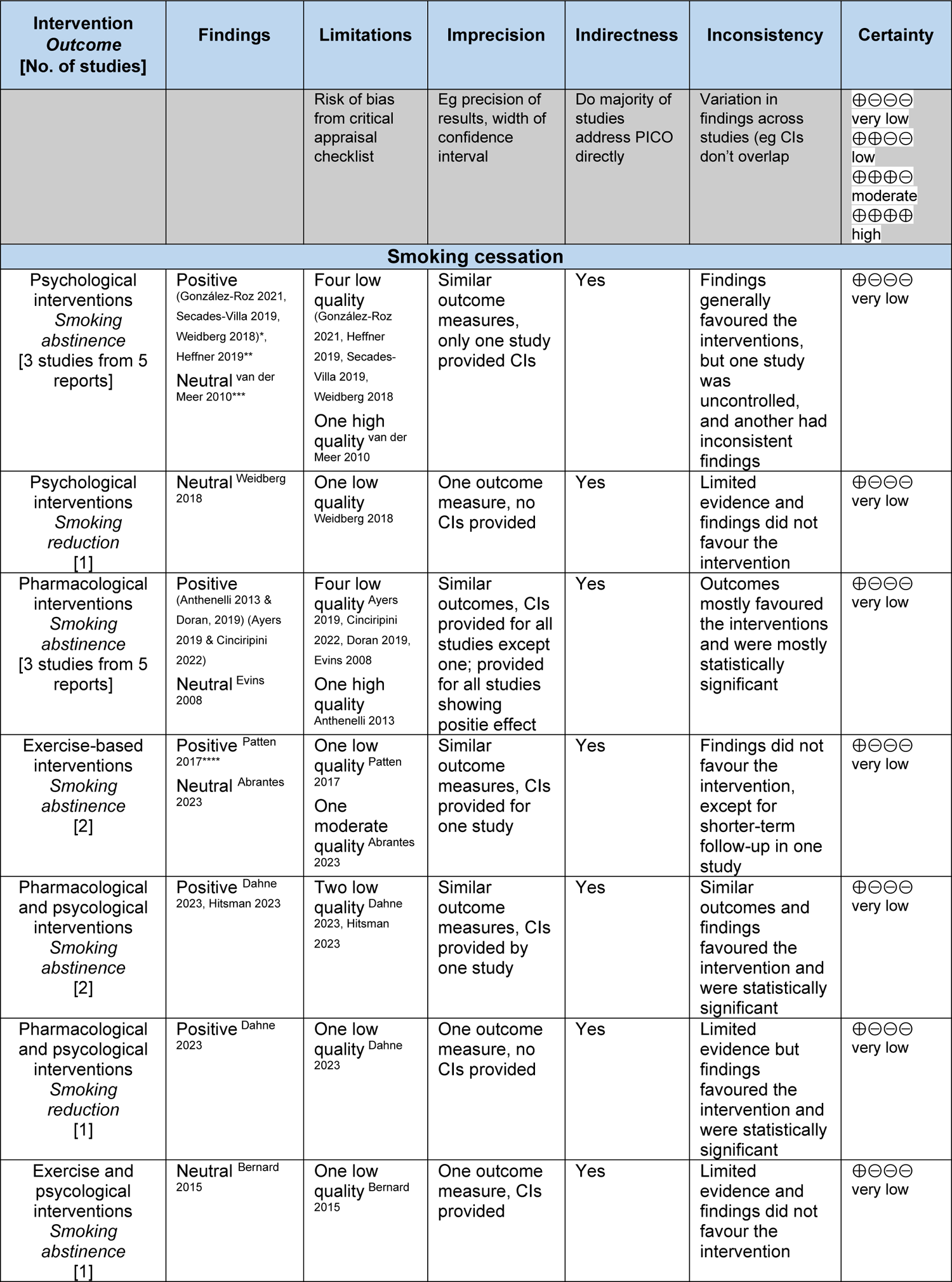
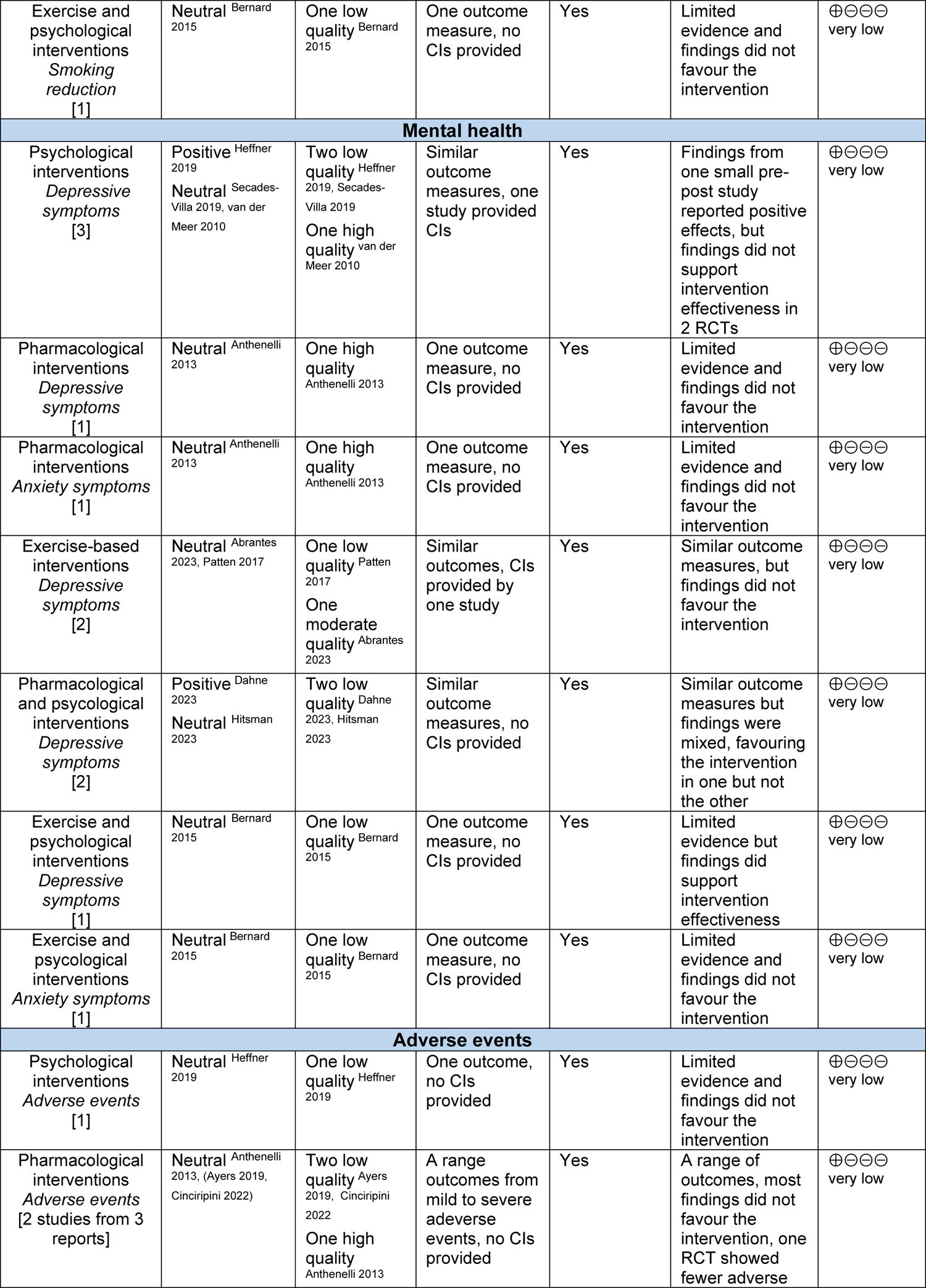
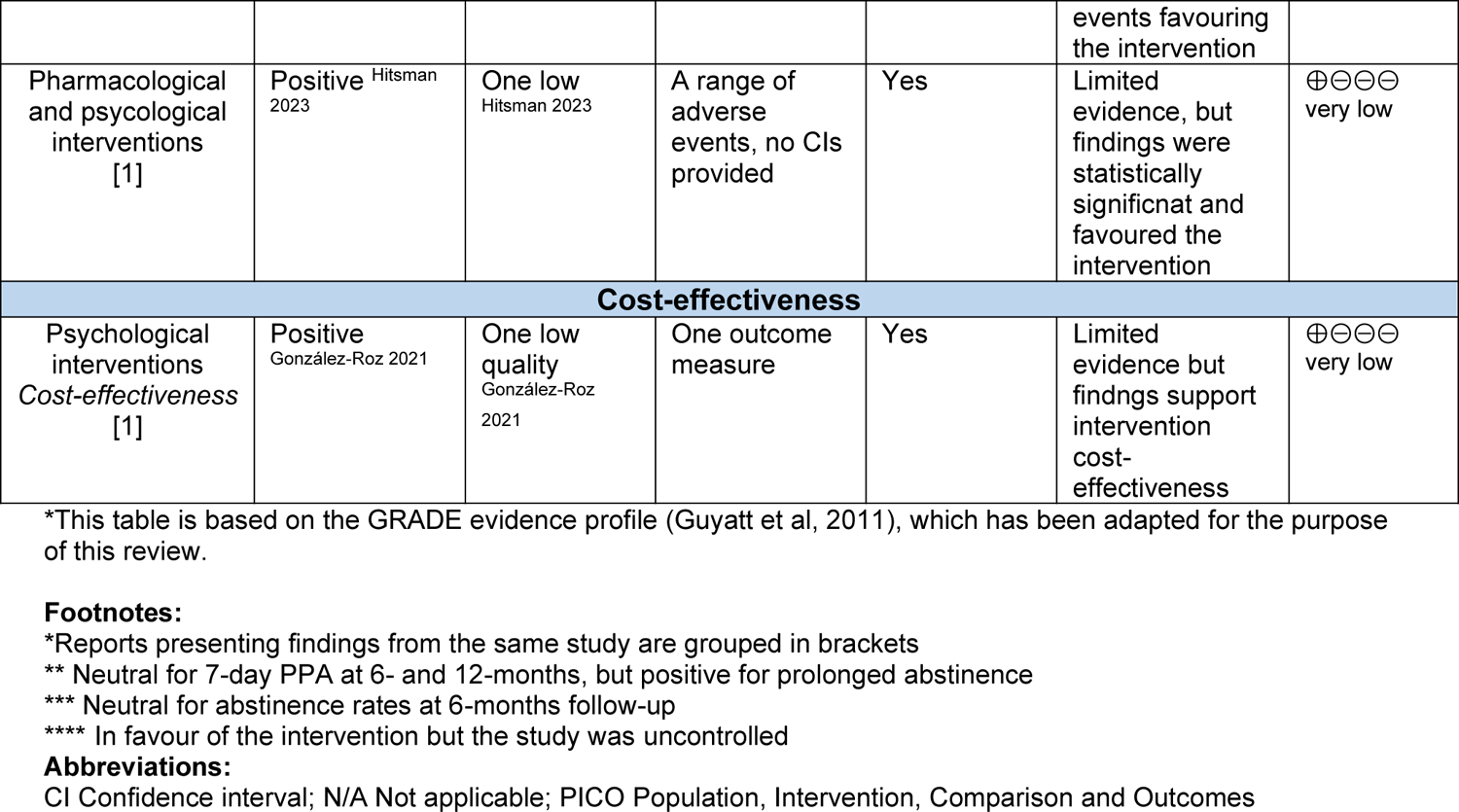
Summary of the evidence*.

Outcome measures identified included smoking abstinence-related outcomes; 7-day point prevalence abstinence (PPA), quit rates, continuous or prolonged abstinence, smoking reduction outcomes; quit attempts, number of cigarettes smoked per day, and cigarette demand, anxiety or depressive symptoms, adverse events, and cost-effectiveness. Data collection methods included self-reported smoking and abstinence rates, validated questionnaires, and tests for biochemical verification were performed in all studies except two (Dahne et al, 2012; van der Meer et al, 2010).

The methodological quality of included studies was assessed using the appropriate Joanna Briggs Institute (JBI) critical appraisal tool for RCTs or quasi-experimental studies. All reports were judged to have some risk of bias, often because of poor reporting of methods. Of the 14 RCT reports, 11 were considered low quality, one was considered moderate quality, and two were considered high quality. Concerns which increased the risk of bias of RCTs included uncertainty about measurement reliability in 11 reports, no description or analysis of loss to follow-up in 11 reports, absence of power calculations stated in 11 reports, and no details about blinding of assessors in 10 reports. The quasi-experimental uncontrolled before and after pilot study was considered low quality due to the absence of a control group and power calculation, therefore it was unclear whether it was sufficiently powered to measure intervention effectiveness. Further details of the quality appraisal can be found in section 6.3.

Multiple reports presented findings from three studies (Anthenelli et al, 2013; Anthenelli et al, 2016; Weidberg et al, 2018), which were included because they provided additional, unique results relevant to the research question. Reports of the same study were identified by trial registration numbers and grouped accordingly. Doran et al (2019) conducted a secondary analysis of the international study reported by Anthenelli et al (2013) (trial number: NCT01078298). Both Ayers et al (2019) and Cinciripini et al (2022) were secondary analyses of the international Evaluating Adverse Events in a Global Smoking Cessation Study (EAGLES) (NCT01456936) (Anthenelli et al, 2016). However, Ayers et al (2019) compares the findings for participants with and without anxiety disorders, whereas Cinciripini et al (2022), compared the findings for participants with and without depression. Anthenelli et al (2016) was not eligible for inclusion because smoking cessation outcomes were not reported specifically for participants with either anxiety or depression. González-Roz et al (2021), Weidberg et al (2018), and Secades-Villa et al (2019), all report findings from the same Spanish study (NCT03163056), although Weidberg et al (2018) report findings from a smaller sample of participants. For the synthesis and summary of study findings (Table 3), reports of the same study have been grouped together.

### 2.2 Impact of psychological interventions

Five reports of three studies (González-Roz et al, 2021; Heffner et al, 2019; Secades-Villa et al, 2019; van der Meer et al, 2010; Weidberg et al, 2018) were examining the effect of psychological interventions for improving smoking cessation and mental health outcomes in smokers with depression. Three of the five reports (González-Roz et al, 2021; Secades-Villa et al, 2019; Weidberg et al, 2018) were analyses of the same smoking cessation study conducted in Spain. The remaining studies were conducted in the Netherlands (van der Meer et al, 2010), and USA (Heffner et al, 2019). All studies were RCTs except for one quasi-experimental before and after pilot study (Heffner et al, 2019). One RCT was considered high quality (van Der Meer et al, 2010), the remaining studies were considered low quality. All studies focused on participants with depression. The findings of studies evaluating the effectiveness of psychological smoking cessation interventions was mixed.

#### 2.2.1 Contingency management

Weidberg et al (2018), Secades-Villa et al, (2019), and González-Roz et al, (2021), all reported findings from the same study where both groups received CBT combined with behavioural activation, and the intervention group received additional contingency management comprising of a voucher-based abstinence reinforcement element. Both interventions were implemented in a group-based format of a maximum of four patients over an 8-week period. Individuals received one therapy session per week and were also asked to attend midweek sessions (“b” sessions) to collect biochemical measures.

##### Smoking abstinence

- Weidberg et al, (2018), assessed smoking status by measuring urine cotinine values, and results showed they were **lower among participants who received CBT with behavioural activation and contingency management compared to those who received only CBT with behavioural activation. Significant differences by group were found in sessions 2b, 3, 3b, 4, 4b, 5 and 6 (all p values <0.05)**. The percentage of participants presenting cotinine levels <80 ng/ml was higher in the CBT with behavioural activation and contingency management group than in the group without contingency management, although differences were only significant in session 5 (p=0.038).
- **Secades-Villa et al (2019) reported that** the odds of a point prevalence abstinence in the group receiving CBT combined with behavioural activation and contingency management were 2.42 times higher [β =0.88, standard error (SE)=0.42, p=0.036, Yule’s Q= 0.42] than that of the control group at 1-month follow-up**. In addition,** continuous abstinence was significantly higher at each time-point up to 6-months follow-up in the group receiving CBT combined with behavioural activation and contingency management (all p values<0.05).
- González-Roz et al (2021) reported that CBT with behavioural activation and contingency management (53.3%; 32/60) was more effective in facilitating 1-year abstinence compared with treatment without contingency management (23.3%; 14/60), however significance values were not reported. **The odds of a favourable response in terms of point prevalence were 1.16 (SE= 0.067, p=0.010) times higher in CBT with behavioural activation and contingency management participants**. Days of **continuous abstinence ranged from 0 to 418 days (CBT with behavioural activation = 97.14 days [SD = 160.70] vs. CBT with behavioural** activation and contingency management = 187.28 days [SD = 182.35], p=0.013), and weeks of longest duration of abstinence were also greater in patients allocated to CBT with behavioural activation and contingency management (25.94 weeks [SD = 24.27] vs. 13.97 weeks [SD = 20.61], p=0.004, d=0.53).

##### Smoking reduction

- Weidberg et al (2018) measured smoking demand through a cigarette purchase task at the end of treatment. There was no significant effect of treatment condition on cigarette demand (Model D: all p values ≤0.32), suggesting that the differential effect of the contingency management component was not significant for this outcome.

##### Mental health

- Secades-Villa et al (2019) measured change in depressive symptoms using the BDI-II scale over a 6-month period. Depressive symptoms significantly reduced amongst all participants, F(5,116) = 57.19, p<0.0001, but there was no significant difference between participants who received the addition of contingency management compared to participants in the control who did not, F(1,111)= 0.53, p=0.4665.

##### Cost-effectiveness

- González-Roz et al (2021) estimated the incremental cost-effectiveness of adding contingency management in relation to abstinence outcomes for a CBT and behavioural activation intervention. **The average cost per patient was €208.85 (US$236.57) for CBT with behavioural activation, and €410.64 (US$465.14) for CBT+BA+CM, p<0.001. The incremental cost of using contingency management to enhance 1-year abstinence by one extra longest duration of abstinence week was €18 (US$20.39) (95% CI 17.75 to 18.25)**.

#### 2.2.2 Behavioural activation & smoking cessation mobile application

Heffner et al (2019) described the development and preliminary evaluation of Actify!-a Behavioural Activation Treatment for Depression (BAT-D) smoking cessation mobile health app (native app on an iOS Apple device) for depressed smokers in a pilot study. Participants received daily messages for 6-weeks, which increased in frequency on and around the quit date.

##### Smoking abstinence

- The findings reported by Heffner et al (2019) showed that the 7-day point prevalence abstinence (PPA) rate at 6-week follow-up was 31% (5/16), and the 30-day PPA at 6-week follow-up fell to 19% (3/16). However, no inferential statistics were reported to support these findings, and the study did not appear to have been powered to detect significant differences.

##### Mental health

- Heffner et al (2019) also measured change in depressive symptoms using the PHQ-9 scale. After the 6-week intervention, **depression scores significantly reduced from baseline (mean –4.5, 95% CI –7.7 to –1.3, p=0.01), which was also a clinically significant change from moderate to mild symptoms**.

##### Adverse events

- Heffner et al (2019) was the only study of a behavioural psychological smoking cessation intervention to record adverse events. During the 6-week intervention, no adverse events were reported by participants from either group.

#### 2.2.3 Mood management

van der Meer et al (2010) investigated whether the mood management component as an adjunct to the Dutch national smoking cessation Quitline (telephone smoking cessation counselling) improved smoking abstinence rates and prevented recurrence of depressive symptoms. The mood management intervention consisted of a tailored self-help mood management manual, as well as two additional counselling sessions, whereas the control group received eight telephone counselling sessions.

##### Smoking abstinence

- Results showed that **prolonged abstinence rates at 6- and 12-month follow-up were significantly higher for participants in the experimental group, 30.5% and 23.9%, respectively, vs 22.3% and 14.0%. At 6-months: OR 1.60 (95% CI 1.06 to 2.42) and at 12-months: OR 1.96 (95% CI 1.22 to 3.14)**. However, although 7-day PPA rates at 6- and 12-month follow-up were higher for participants in the experimental group, 37.4% and 27.6%, respectively, vs 31.0% and 24.0%, this was not statistically significant at 6-months: OR 1.38 (95% CI 0.94 to 2.02) or 12-months: OR 1.23 (95% CI 0.81 to 1.86).

##### Mental health

- van der Meer et al (2010) measured changes in depressive symptoms using the CES-D scale. There were no significant differences between groups at 6- and 12-months; scores reduced by a mean of 1.1 and 0.6 respectively for participants in the intervention group, and by 2.0 and 0.1 in the control (p>0.05).

#### 2.2.4 Bottom line results for psychological interventions

Overall, there is very low certainty evidence that suggests psychological interventions may be effective for improving smoking cessation in people with depression. Certainty is limited because of the small number of studies, which were mostly of low-quality. The only study to suggest psychological interventions improve mental health symptoms, was a small, low-quality quasi-experimental pilot study without a control group. There was no evidence to suggest psychological interventions result in fewer adverse events, however, none of the interventions appeared to cause greater harm. There was limited evidence to suggest that psychological smoking cessation interventions may be cost-effective. However, as these outcomes were reported by single studies which were of low quality, firm conclusions cannot be made. The evidence supporting the use of psychological interventions for improving mental health outcomes in smokers with depression appears inconsistent.

### 2.3 Impact of pharmacological interventions

Five reports were examining three different studies exploring the effect of pharmacological interventions on smoking cessation and mental health outcomes (Anthenelli et al, 2013; Ayers et al, 2019; Cinciripini et al, 2022; Doran et al, 2019; Evins et al, 2008). Four reports were of two studies two studies conducted internationally across the EU and USA (Anthenelli et al, 2013; Anthenelli et al, 2016), and one study was conducted in the USA alone (Evins et al, 2008). One study was considered high quality (Anthenelli et al, 2013), while the remaining studies were considered low quality. Four reports focused on participants with depression, and one on participants with anxiety (Ayers et al, 2019). Findings in relation to the effectiveness of the interventions appear mixed.

#### 2.3.1 Varenicline

The effect of Varenicline on smoking cessation was assessed by two studies reported by four reports. Anthenelli et al (2013) and Doran et al (2019) both evaluated smoking abstinence and changes in mood and anxiety levels in smokers with depression treated with Varenicline versus placebo. All participants received smoking cessation counselling, however the effects of Varenicline were compared to a control group which received a placebo. A dose of 1mg of Varenicline was taken twice daily, or placebo, over 12-weeks.

Ayers et al (2019) and Cinciripini et al (2022), reported findings from the EAGLES trial (Anthenelli et al, 2016) comparing the safety and efficacy of Varenicline, Bupropion, NRT, and placebo over 12-weeks. The study administered a dose of 1mg of Varenicline twice daily, 150mg of Bupropion twice daily, or transdermal nicotine patch (NRT) at a dose of 21 mg/day with taper, plus weekly cessation counselling for 12 weeks. The outcomes for each pharmacological intervention were compared with participants in a control group who received smoking cessation counselling and a placebo.

##### Smoking abstinence

- Results reported by Anthenelli et al (2013) showed **7-day point prevalence abstinence (PPA) rates were higher for participants who received Varenicline compared to placebo across all time points; week 12; 46.1% vs. 20.1% (OR 3.82, 95% CI 2.53 to 5.78, p<0.001), week 24; 31.3% vs. 18.2% (OR 2.16 95% CI, 1.40 to 3.33, p<0.001), and week 52; 28.5% vs. 17.5% (OR 1.98, 95% CI 1.28 to 3.08, p=0.002)**. In addition, **continuous abstinence rates were also higher at every time point for those in the Varenicline group compared to placebo; weeks 9 to 12; 35.9% vs. 15.6% (OR 3.35, 95% CI 2.16 to 5.21, p<0.001), weeks 9 to 24; 25.0% vs. 12.3% (OR 2.53, 95% CI 1.56 to 4.10, p<0.001), and weeks 9 to 52; 20.3% vs. 10.4% (OR 2.36 95% CI 1.40 to 3.98, p<0.001)**.
- Doran et al (2022) was a secondary analysis of Anthenelli et al (2013) and sought to assess the impact of baseline depressive symptoms on abstinence. The results showed at the mean level of baseline depressive symptoms, **participants who received Varenicline were nearly four times more likely to achieve continuous abstinence than those who received the placebo at weeks 9-12 (OR 3.91, 95% CI 2.54% to 6.34%, p<0.001)**. In addition, higher baseline symptoms of depression were associated with significantly lower likelihood of abstinence for the placebo group (OR 0.91, 95% CI 0.85% to 0.97%, p=0.004), but not the Varenicline group (OR 0.99, 95% CI 0.95% to 1.03%, p=0.568). For PPA outcomes, between 13-52 weeks measurement, significant treatment (z= 5.88, p<0.001) and treatment by time (z = −3.28, p=0.001) effects indicated an initial advantage of Varenicline which faded over the 52-weeks, however, PPA rates remained greater in the Varenicline group compared to placebo throughout.
- Ayers et al (2019) reported findings from a subgroup analysis of participants with anxiety from the EAGLES trial (Anthenelli et al, 2016). The results showed **7-day PPA rates were higher for participants with anxiety who received Varenicline compared to placebo at week 12 (OR 4.65, 95% CI 1.95 to 1.11), and week 24** (OR 3.13, 95% CI 1.28 to 7.62). Seven-day PPA rates were also higher for participants with anxiety who received Varenicline compared to Bupropion (OR 2.80, 95% CI 1.22 to 6.44), and NRT (OR 2.32, 95% CI 1.04 to 5.17) at week 12, but there was no significant difference at week 24. **Continuous abstinence rates at weeks 9-12 were also greater for participants with anxiety who received Varenicline compared to placebo (OR 4.53, 95% CI 1.20 to 17.10)**. There were no significant effects in favour of Bupropion or NRT in any comparisons for participants with anxiety.
- Cinciripini et al (2022) reported findings from a subgroup analysis of participants with depression versus those without a psychiatric disorder from the EAGLES trial (Anthenelli et al, 2016). The results showed that **compared to placebo, continuous abstinence rates were higher for participants with depression who received Varenicline at weeks 9-12 (OR 3.004, 95% CI 2.081 to 4.336) and weeks 9-24 (OR** 2.396, 95% CI 1.561 to 3.678). Compared to Bupropion, odds of continuous **abstinence were greater for Varenicline at weeks 9-12 (OR 1.551, 95% CI 1.118 to 2.152)** but not at weeks 9-24 (OR 1.295, 95% CI 0.888 to 1.890). **compared to NRT, odds of continuous abstinence were greater for Varenicline at weeks 9-12 (OR 1.589, 95% CI 1.146 to 2.202) and weeks 9-24 (OR 1.574, 95% CI 1.066 to** 2.325).

##### Mental health

- Anthenelli et al (2013) measured changes in anxiety and depression symptoms amongst participants and found no significant differences between groups. Both groups saw similar changes which trended toward a slight improvement, but values were not given within the study. Furthermore, there were no clinically relevant differences between groups in suicidal ideation or behaviour, and there was no worsening of anxiety or depression.

##### Adverse events

- Anthenelli et al (2013) reported treatment-emergent adverse events, defined as events which occurred either during treatment, or up to 30-days post-treatment. Adverse events included sleep, anxiety, depression and mood, personality and behaviour, physical activity, and suicide and self-harm events. Adverse events occurred in 72.3% of Varenicline participants and 66.9% of placebo participants, most were rated as mild or moderate, the most frequent adverse event was nausea (Varenicline, 27.0%; placebo, 10.4%). Adverse events led to treatment discontinuation in 6.3% and 7.8% of Varenicline and placebo participants, respectively, however, inferential statistics were not reported within the study to determine whether these differences were statistically significant.

#### 2.3.2 Bupropion

Evins et al (2008) sought to determine if Bupropion improved abstinence rates when added to transdermal NRT and group CBT. The intervention was delivered over 13-weeks, and comparison were made with a control consisting of the same NRT and CBT with a placebo. Cinciripini et al (2022), compared the findings for participants who received Bupropion compared to those who received the placebo control in the EAGLES trial (Anthenelli et al, 2016). The comparisons were made in a subgroup analysis of participants with depression versus those without a psychiatric disorder.

##### Smoking abstinence

- Results reported by Evins et al (2008) showed that the 7-day PPA rate was higher for participants who received Bupropion in addition to CBT and NRT compared to the control group, but the difference was not statistically significant; 36% (37/97), vs 31% (32/102), p=0.49.
- Results reported by Cinciripini et al (2022) showed that **compared to placebo, odds of continuous abstinence were also greater for participants with depression who received Bupropion at weeks 9-12 (OR 1.936, 95% CI 1.321 to 2.838) and** weeks 9-24 (OR 1.85 95% CI 1.188 to 2.881),

#### 2.3.3 Nicotine replacement therapy

As well as comparing the effect of Varenicline and Bupropion compared to placebo, Cinciripini et al (2022) also reported the effect of NRT compared to placebo in a sub-group analysis of participants with depression versus those without a psychiatric disorder from the EAGLES trial (Anthenelli et al, 2016).

##### Smoking abstinence

- The results reported by Cinciripini et al (2022) showed that **compared to placebo, odds of continuous abstinence for participants who received NRT were only significantly greater at weeks 9-12 (OR 1.891, 95% CI 1.291 to 2.770), but not** weeks 9-24 (OR 1.522, 95% CI 0.967 to 2.396).

##### Adverse events

- Anthenelli et al (2013) reported treatment-emergent adverse events as part of the EAGLES trial, defined as events which occurred either during treatment, or up to 30-days post-treatment. Adverse events included sleep, anxiety, depression and mood, personality and behaviour, physical activity, and suicide and self-harm events. Adverse events occurred in 72.3% of Varenicline participants and 66.9% of placebo participants, most were rated as mild or moderate, the most frequent adverse event was nausea (Varenicline, 27.0%; placebo, 10.4%). Adverse events led to treatment discontinuation in 6.3% and 7.8% of Varenicline and placebo participants respectively, however, inferential statistics were not reported within the study to determine whether these differences were statistically significant.
- Ayers et al (2019) also reported adverse events from the EAGLES trial (Anthenelli et al, 2016) and the results showed that participants with generalised anxiety disorder had an increased incidence of moderate-to severe neuropsychiatric adverse events compared to non-psychiatric smokers (5.4% vs 2.1%, p=0.0012), but there were no significant differences in moderate to severe neuropsychiatric adverse events between treatment groups.
- Cinciripini et al (2022) also reported adverse events from the EAGLES trial (Anthenelli et al, 2016), and the results showed that the risk of neuropsychiatric adverse events did not differ by medication in participants with depression (p>0.05).

##### Bottom line results for pharmacological interventions

There is very low certainty evidence to suggest that pharmacological interventions may be effective at improving smoking cessation outcomes, although the evidence was mostly low quality. Most of the available evidence focuses on interventions for people with depression, as only one study analysed the effects in participants with anxiety disorders. Each study that evaluated the impact of interventions which used Varenicline reported statistically significant improvements in cessation outcomes, and some of the evidence suggests Bupropion and NRT were also effective, although the evidence was mixed, and particularly limited for NRT.

The evidence supporting the use of pharmacological interventions for improving mental health symptoms in smokers is lacking. It is worth noting that this outcome was examined by only one study. In addition, there was no evidence to suggest that pharmacological interventions resulted in fewer adverse events, but neither was there any evidence to suggest they cause greater harm.

### 2.4 Impact of aerobic exercise-based interventions

Two studies assessed the impact of aerobic exercises of varying intensities on smoking cessation and mental health (Abrantes et al, 2023; Patten et al, 2017). Both studies were RCTs conducted in the USA and included participants with depression. One study was considered high quality (Abrantes et al, 2023), while the other was considered low quality. Findings in relation to the impact of the interventions appear mixed.

#### Smoking abstinence

- Abrantes et al (2023) tested the efficacy of a 12-week moderate intensity aerobic exercise intervention compared to a health education contact as an adjunct to standard smoking cessation treatment in individuals with depression. Study findings reported no significant differences in 7-day point prevalence abstinence (PPA) between groups at follow up (OR 0.78; 95% CI 0.32 to 1.94; p=0.60).
- Patten et al (2017) conducted a pilot study to evaluate the potential role of supervised vigorous exercise as a smoking cessation intervention for depressed female smokers. Study participants were randomly assigned to 12 weeks of thrice weekly exercise or health education, with both groups receiving behavioural smoking cessation counselling and NRT. The study found that **after 12 weeks, smoking abstinence rates were significantly higher for participants who received the exercise intervention compared to the health education group (73% [11/15]) vs 33% [5/15]); χ2 =4.821, df= 1, p=0.028)**. At 6-months follow-up, there was no statistically significant differences in abstinence rates between groups (27% [4/15] for the exercise condition vs.40% [6/15] for health education; χ2= .600, df= 1, p=0.439).

#### Mental health

- Abrantes et al (2023) measured changes in depressive symptoms using the CES-D scale and reported no significant differences between participants who received the aerobic exercise intervention and those who received health education (b = −0.84; 95% CI −3.25 to 1.57; p=0.49).
- Patten et al (2017) measured change in depressive symptoms using the PHQ-9 scale and reported no significant differences at 12 weeks between groups receiving either the exercise intervention or health education (p=0.90).

#### 2.4.2 Bottom line results for exercise-based interventions

There is very low certainty evidence that suggests exercise interventions as adjuncts to standard smoking cessation treatments in individuals with depression may not be effective. Only two studies evaluating exercise interventions were identified, both of which had contrasting findings in relation to smoking abstinence, and neither study was of high quality. Exercise interventions did not demonstrate any positive impact on alleviating depressive symptoms.

### 2.5 Impact of multicomponent interventions comprising of pharmacological and psychological components

Two studies investigated the effect of a combination of pharmacological and psychological interventions on smoking cessation and mental health outcomes (Dahne et al, 2023; Hitsman et al, 2023). Both studies were RCTs conducted in the USA and recruited participants with depression. Findings appear to be in support of the interventions.

#### 2.5.1 Behavioural activation mobile phone application with nicotine replacement therapy

Dahne et al (2023) assessed the efficacy of the Goal2Quit intervention, a behavioural activation-based self-directed mobile phone application for treating depression and quitting smoking. This was combined with a two-week course of NRT, including 14mg nicotine patches and 4mg nicotine lozenges. The control group received digital copy of the National Cancer Institute’s “Clearing the Air: Quit Smoking Today” booklet.

##### Smoking abstinence

- The study findings reported by Dahne et al (2023) showed that **both 7-day point prevalence abstinence (PPA) and floating abstinence were significantly higher for Goal2Quit participants at 4-8- and 12-weeks follow-up compared to control**. **At 12-weeks**, **the 7-day PPA rate was 16% vs 2% (p=0.02), and the floating abstinence rate was 24% vs 4% (p=0.003)**.

##### Smoking reduction

- Dahne et al (2023) measured self-reported 24-hour quite attempts, and number of cigarettes smoked per day. **Goal2Quit participants were more likely to have made a 24-hour quit attempt during the first 4-(17% vs 2%; p=0.01), 8-(24% vs 4%; p=0.003), and across all 12-(29% vs 6%; p=0.002) weeks follow-up**. Participants in the **Goal2Quit group also reported smoking less cigarettes per day than the treatment as usual group (difference of mean 1.97, SE 0.93 CPD less; p=0.03)**.

##### Mental health

- Dahne et al (2013) measured change in depressive symptoms using the BDI-II scale and reported that depressive symptoms decreased in both groups, but **the Goal2Quit group reported significantly lower average depressive symptoms compared to the treatment as usual group (difference of mean 3.72, SE 1.37 points less for Goal2Quit; p=0.01)**. In comparison to 14 participants from the treatment as usual group, 23 Goal2Quit participants reported worsening mood over the course of the treatment, but four participants of the Goal2Quit group endorsed suicidality, compared to seven in the treatment as usual group. Inferential statistics were not given to determine whether they were statistically significant.

#### 2.5.2 Behavioural activation with Varenicline

Hitsman et al (2023) measured the efficacy and safety of combining 12-weeks of behavioural activation for smoking cessation and Varenicline for smokers with depression. There were four comparator groups: behavioural activation for smoking cessation with Varenicline or a placebo, and standard behavioural treatment and Varenicline or a placebo.

##### Smoking abstinence

- Results reported by Hitsman et al (2023) showed **that abstinence rates at 27-weeks were higher in participants in the two groups who received Varenicline compared to placebo, rate ratio (RR) 2.16 (95% CI 1.08 to 4.30, p=0.0277)**. Continuous abstinence rates were similar between groups, however, prolonged abstinence over 14- and 27-weeks favoured the Varenicline arms, although significance values were not reported.

##### Mental health

- Hitsman et al (2023) measured changes in depressive symptoms using the BDI-II scale, and results showed no statistically significant differences in depression levels among treatment arms at any of the follow-up timepoints (p>0.05).

##### Adverse events

- Hitsman et al (2023) reported adverse events and resulted showed that **at 6 weeks, statistically significant differences were reported with the placebo group experiencing more sleep problems (p=0.035) than those receiving Varenicline. At 14-weeks statistically significant differences were also reported with the placebo group experiencing increased rates of dry mouth (p=0.009) and anxiety (p=0.023**).

#### 2.5.3 Bottom line results for multicomponent interventions (pharmacological and psychological)

There is very low certainty evidence to suggest that multicomponent interventions comprising of pharmacological and psychological components may be effective at improving smoking cessation outcomes in people with depression. However, the evidence supporting the use of these interventions in improving depressive symptoms and adverse events is limited, in particular for adverse events which was only reported by a single study.

### 2.6 Impact of multicomponent interventions comprising of exercise and psychological intervention components

One study examined the use of a multicomponent intervention comprising of an exercise and behavioural component (Bernard et al, 2015). This study was a pilot RCT conducted in France and focused on smokers with depression. It was considered low quality, and the findings did not favour the intervention.

#### Smoking abstinence

- Bernard et al (2015) sought to estimate the effects of adding exercise and counselling to standard smoking cessation treatment (consisting of NRT or Varenicline), for smokers with depressive disorders, compared to a health education control. The study results showed that although at 12-weeks from baseline continuous abstinence rates were higher in the experimental group (48.5% (17/35) vs 28.5% (10/35); χ2(7) = 2.95, p=0.08), there was no statistically significant difference between groups at any time-point, and no significant differences in continuous abstinence rates were detected.

#### Smoking reduction

- Bernard et al (2015) measured the number of self-reported cigarettes smoked per day and found no significant between-group differences in the number of cigarettes smoked per day between participants who received exercise and counselling intervention, compared to participants in the control group.

#### Mental health

- Bernard et al (2015) measured change in depressive and anxiety symptoms at weeks 8, 12, 24 and 52 using the HADS scale, but there were no significant differences in depressive or anxiety symptoms at any time point.

#### 2.6.2 Bottom line results for multicomponent interventions (exercise and psychological)

There is very low certainty evidence that suggest multicomponent interventions comprising of exercise and psychological interventions may not be effective at improving smoking cessation in individuals with depression. However, the evidence is limited as only one, low quality study (a pilot RCT) was identified, and this did not show any significant differences for smoking cessation or mental health outcomes.

## 3. DISCUSSION

### 3.1 Summary of the findings

This rapid review aimed to identify and synthesise the evidence for the effectiveness of smoking cessation interventions in people with anxiety or depression living in the community. There is some, mostly low quality, evidence to suggest that smoking cessation interventions may be effective at improving a range of smoking cessation and mental health outcomes in people with depression. Only one study was identified assessing smoking cessation interventions in people with anxiety meaning conclusions about the evidence of effectiveness cannot be drawn. Psychological interventions may increase abstinence in smokers with depression, but results were mixed regarding their impact on mental health outcomes. Pharmacological interventions, particularly Varenicline, appear effective for smoking cessation in people with depression and anxiety. However, evidence for Bupropion and NRT is limited and mixed. Findings generally did not support the effectiveness of exercise interventions for smoking cessation, and no sustained benefits or improvements in depressive symptoms were identified. Although the evidence was limited, none of the studies suggested that the interventions caused greater harm than the smoking cessation treatment received by the control group. Delivering interventions as part of a multicomponent approach may be beneficial, but methodological limitations and variation in intervention types and outcome measures across studies compromise the applicability of these findings. Further research with rigorous methodologies is needed to provide clearer guidance on effective smoking cessation strategies for this population.

### 3.2 Strengths and limitations of the available evidence

A strength of the available evidence is the identification of a wide range of smoking cessation interventions. However, the considerable heterogeneity across studies makes it difficult to compare interventions directly and limits the overall certainty of the evidence base.

Variation in recruitment strategies raises questions about the similarities of participants across the included studies. Furthermore, inconsistency in diagnostic tools and cut-offs scores for inclusion hinders comparisons of depression severity and intervention effectiveness across studies.

The rapid review identified limited, low-quality evidence for smoking cessation interventions in individuals with anxiety or depression. Most studies lacked methodological rigor and were poorly reported. Only two RCTs were deemed high quality, while industry funding and involvement in five studies raised concerns about potential bias. Although 11 studies were identified, three were small pilot studies. These pilot studies, while potentially valuable for informing future research directions, were not powered sufficiently to detect intervention effectiveness due to their limited sample sizes. Additionally, seven reports were secondary analyses of three primary studies. Although secondary analyses may provide data from original studies with greater relevance to the research question (e.g., depression sub-group analysis), they may not be sufficiently powered and there is potential for overlap of participants across reports. If multiple reports draw data from the same original study population, this can inflate the overall sample size but not necessarily represent new data or a broader participant pool. Furthermore, data is limited by the original studies’ research questions and data collection methods. These limitations might mean the data wasn’t specifically collected to answer questions related to smoking cessation interventions for individuals with anxiety or depression.

Several studies included in this rapid review involved interventions being assessed as an addition to existing smoking cessation treatment which was received by all participants. It is important to consider that the relative effect of the intervention compared to the control may be diminished due to the underlying smoking cessation treatment received by all participants. While examining the effect of an intervention as an addition to existing treatment allows for an assessment of specific intervention components, the underlying treatment received by all participants likely affects the overall findings.

Several evidence gaps were identified. Notably, none of the included studies were conducted within the UK. This absence of recent UK-based research limits our understanding of how these interventions might translate to the specific healthcare system and cultural context of the Wales. Although no published UK studies met our criteria, a protocol for a UK-based smoking cessation intervention for smokers with depression was identified. This study, currently under review for publication in the Addiction journal (ISRCTN99531779; https://doi.org/10.1186/ISRCTN99531779), is especially pertinent to this rapid review as it explores smoking cessation support integrated into routine psychological care within the UK. Additionally, the review identified gaps relating to barriers and facilitators to smoking cessation interventions, and the populations studied. Only one study focused on participants with anxiety disorders. No studies reported co-existing mental health conditions, which would have provided a more nuanced understanding of intervention effectiveness within this population. These limitations suggest a need for caution when generalising the findings to broader populations.

This rapid review found no evidence on equity factors and intersectionality. Included studies did not consider how socioeconomic factors, ethnicity, or social capital affect access and success of interventions. Understanding these complexities is essential for developing inclusive and equitable smoking cessation programs.

Additionally, the rapid review found no studies investigating harm reduction approaches, and limited research reporting interventions applied at critical touchpoints within healthcare systems and smoking cessation services (where targeted interventions could reach the review’s included population), or interventions that integrate with existing healthcare pathways or mental health services.

This rapid review identified no studies evaluating the effectiveness and safety of e-cigarettes as a smoking cessation or harm reduction tool for people with anxiety or depression. This highlights an absence of research on e-cigarettes as a smoking cessation or harm reduction aid in our population of interest. E-cigarettes have become a popular alternative to traditional cigarettes, potentially offering a route to harm reduction or smoking cessation for smokers with depression or anxiety. Despite their potential benefits, the rising prevalence of e-cigarette use among non-smokers, particularly children and young people, raises concerns which requires further investigation.

Only one study included in this rapid review analysed intervention cost-effectiveness, which investigated the one-year cost-effectiveness of adding contingency management to cognitive-behavioural treatment and behavioural activation for quitting smoking in smokers with depression in Spain. Despite reporting positive outcomes however, we cannot comprehensively assess the cost-effectiveness of such interventions due to insufficient evidence.

### 3.3 Strengths and limitations of this Rapid Review

The studies included in this rapid review were systematically identified through an extensive search of electronic databases and grey literature. The review was also based on an abbreviated systematic review approach, following established guidelines for conducting rapid reviews in an attempt to capture all relevant publications with minimal risk of bias in a timely manner.

Due to the short timeframe to complete this rapid review, as well as the large number of references initially identified from literature searches, most reports were screened by a single reviewer at title and abstract level. However, 558 references (14%) were double screened to assess consistency. The remaining stages of the review process included consistency checking by at least one other reviewer, with issues being discussed and resolved within the team.

A key strength of this rapid review is its focus on using rigorous participant selection criteria for identifying relevant studies. Studies were only considered eligible if they recruited participants using validated measurement tools for depression and anxiety, and specific cut-off points, depending on the tool used. This approach enabled us to include people with a clinical diagnosis already within the healthcare system, and those who should be but are not currently in the system. Although these two groups may have different needs and respond differently to smoking cessation interventions, we were able to capture a more comprehensive picture of the effectiveness of these interventions within a population accessing the healthcare system.

However, limitations exist. It is important to note that many studies excluded participants who were currently prescribed medication for depression or anxiety. This exclusion may have inadvertently left out individuals who are currently within the healthcare system and who may have responded differently to the intervention than those included in the study.

We excluded primary studies that investigated intervention effectiveness by comparing those with anxiety or depression with a non-psychiatric group (see Appendix 2). This was because our rapid review focussed on people with anxiety and depression, therefore we wanted to compare effectiveness in this population only. However, including these studies may have provided a more detailed understanding of intervention effectiveness.

Furthermore, the review excluded studies relying solely on self-reported depression, potentially missing individuals who experience symptoms but lack a formal diagnosis – a potential hard-to-reach group likely to have been of interest to our stakeholders. Second, studies focusing on anxiety sensitivity were also excluded. While anxiety sensitivity is often related to anxiety disorders, we excluded studies focused on this population to maintain focus on interventions specifically targeting people with anxiety or depression, ensuring our findings provide clear conclusions relevant to these populations. Finally, the rapid review did not include studies investigating mixed mental health populations unless they provided separate analyses for participants meeting specific criteria for anxiety or depression. These limitations restrict the generalisability of the findings to a broader population struggling with co-occurring mental health challenges and potentially experiencing symptoms that fall outside the strict diagnostic categories employed in the rapid review.

### 3.4 Implications for policy and practice

This rapid review identified a lack of high-quality evidence assessing the effectiveness of smoking cessation interventions for individuals with anxiety or depression. However, despite this, findings identified could help to inform future intervention development and delivery in this underserved population. It is, however, important to consider the context in which the interventions outlined here were evaluated and the uncertainty of the findings as outlined in table 3.

The rapid review identified Varenicline as a potentially effective smoking cessation medication. Varenicline was manufactured by Pfizer, who withdrew it from the market in 2021. More recently, another manufacturer applied for approval from the Medicines and Healthcare products Regulatory Agency (MHRA) for a generic Varenicline. Additionally, as of 09/07/2024, a different treatment with similar action to Varenicline called Cytosine, has been recommended for use in the general population by the All Wales Medicine Strategy Group, and is expected to be available on prescription within two months.

### 3.5 Implications for future research

Our rapid review identified a paucity of UK-based research in this area, indicating a need for future investigations within the UK to comprehensively assess the effectiveness and generalisability of interventions, particularly within the Welsh context.

While several RCTs were identified, the risk of bias assessment revealed considerable methodological limitations. Future research should prioritise conducting high-quality research with robust methodologies to minimise bias and improve the overall certainty of the evidence base.

There is a notable gap in research on interventions for individuals with predominant anxiety disorders. Future studies should focus on tailored interventions for this population group.

Additionally, our review excluded those self-reporting depression or anxiety in order to identify a population most similar to those engaged with mental health services. Studies which included populations with mixed mental health conditions were also excluded from our review if no sub-group analyses was conducted specifically in participants with either anxiety and/or depression. However, most studies with mixed mental health populations did not perform such analyses. Therefore, future research that includes participants with a range of mental health conditions should conduct analyses to identify the effectiveness of interventions for participants with specific mental health conditions, as it is likely that their needs for support to aid quitting smoking, and the effectiveness of interventions, will vary. This will help to provide greater insight to inform policy for people with specific mental health conditions.

While the rapid review identified various interventions for smoking cessation in individuals with depression, translating efficacy trials into real-world practice remains challenging. Prioritising effectiveness trials within real-world settings is crucial for developing impactful interventions tailored to the Welsh context. In particular, policymakers need evidence-based approaches that can be incorporated into the current system in Wales, enabling them to leverage existing resources.

To strengthen research on smoking cessation interventions for individuals with depression or anxiety, standardised measurement using validated diagnostic tools is essential. Transparent reporting of eligibility criteria will facilitate comparisons to real world settings and build a more robust evidence base. Future research should also include full economic evaluations or cost-effectiveness analyses to provide insights into the financial implications of implementing these interventions within healthcare systems.

### 3.6 Economic considerations*

- The total economic cost of smoking in people with mental health disorders in the UK is estimated at £3.5 billion** per annum. Of this total cost, £1.1 billion** is attributed to smoking-related healthcare treatment, £1.2 billion** to absenteeism and productivity losses and £1.2 billion** to premature mortality (Wu et al., 2015).
- There is limited economic evidence on the impact of implementing smoking cessation interventions for individuals living with depression and/or anxiety.
- Economic modelling evidence suggests tailored smoking cessation interventions combining behavioural and pharmaceutical support for individuals with severe mental illness can be cost-effective compared to usual care. Severe mental illness in this study concerned any mental illness causing significant functional impairment and limits on major life activities (Mattock, Owen & Taylor, 2023).

*This section has been completed by the Centre for Health Economics & Medicines Evaluation (CHEME), Bangor University

** Figures have been inflated to May 2024 prices using the Bank of England Inflation Calculator. These figures assume no difference in observed patterns since the original calculation.

## Abbreviations

Acronym: Full Description

BDI-II: Beck Depression Inventory version 2

CBT: Cognitive behavioural therapy

CES-D: Centre for Epidemiological Studies Depression scale

DSM-IV: Diagnostic and Statistical Manual of Mental Disorders-IV

EAGLES: Evaluating Adverse Events in a Global Smoking Cessation Study

HADS: Hospital Anxiety and Depression scale

ISRCTN: International Standard Randomised Controlled Trial Number

MADRS: Montgomery-Asberg Depression Rating Scale

MHRA: Medicines and Healthcare products Regulatory Agency

MINI: Mini International Neuropsychiatric Interview

NRT: Nicotine replacement therapy

PHQ 8/9: Patient Health Questionnaire versions 8/9

PPA: Point prevalence abstinence

RCT: Randomised controlled trial

## Data Availability

All data produced in the present study are available upon reasonable request to the authors

## RAPID REVIEW METHODS

We searched for primary sources to answer the review question: what is the effectiveness of smoking cessation interventions for people with anxiety and/or depression living in the community.

### 5.1 Eligibility criteria

**Table 4.** Eligibility criteria.

|  | **Inclusion criteria** | **Exclusion criteria** |
| --- | --- | --- |
| **Participants** | Adults aged ≥18 with a current diagnosis, awaiting a diagnosis, or who met the diagnostic criteria of anxiety or depression using a validated screening tool, living in the community who smoke tobacco.<br>Those with another mental health condition, including substance use disorder, will be included if co-occurring with anxiety or depression. | People aged <18 years.<br>Pregnant or breastfeeding women. People with any other mental health condition, if not co-occurring with anxiety or depression.<br>Any inpatients within mental health or other clinical settings. |
| **Settings** | Community, home, primary care, or community mental health team settings. | Any setting which isn't within the community. |
| **Intervention / exposure** | Tobacco smoking cessation interventions or harm reduction approaches delivered to people with anxiety or depression living within the community. This could include:<br>• Interventions that are effective in the general population<br>• Interventions that have been modified to better suit the population of interest<br>• Interventions specifically designed for the population of interest | Interventions or harm reduction approaches which aren't delivered to people with anxiety or depression living within the community. |
| **Control or counter intervention** | No intervention, alternative population, alternative interventions, usual care. |  |
| **Outcome measures** | Any outcome relating to smoking cessation or reduction, anxiety and depression, or cost-effectiveness.<br>Facilitators and barriers if they were included as part of an evaluation. |  |
| **Study design** | Clinical-effectiveness: Primary research studies which use comparative designs.<br>Cost-effectiveness: Full economic evaluations | Primary studies which aren't comparative, qualitative studies, secondary/tertiary research. |
| **Countries** | OECD countries. | Non-OECD countries. |
| **Language of publication** | English language only. | Any other language. |
| **Publication date** | Published between 2007 and 2024. | Published before 2007. |
| **Publication type** | Published and preprint (preprint only if limited published research identified). | Commentaries, editorials, letters, conference abstracts |

### 5.2 Literature search

A search was conducted of the electronic bibliographic databases: Medline, Embase, Scopus, and CINAHL Plus with Full Text. A search for ongoing and completed trials was conducted in Clinicaltrials.gov, and WHO International Clinical Trials Registry Platform (ICTRP). A search for grey literature was conducted of the Action on Smoking and Health (ASH) website. Citation tracking from secondary sources identified during the preliminary stages was also undertaken. All searches were conducted between 15/03/2024 and 21/03/2024. Search concepts and keywords included smoking cessation, stop smoking, smoking reduction, depression, anxiety, and terms to reflect the study designs of interest. Searches were limited to items published after 2007 – the year the smoke free legislation was introduced in Wales, and to those published in the English language.

### 5.3 Study selection process

All studies were uploaded to the systematic reviewing platform Rayyan for screening. For title and abstract screening, titles were screened by a single reviewer. An online random number generator was used to select a sample of 10% (n=387) of all titles, which were screened by a second reviewer to assess consistency with the first reviewer. An additional 171 titles which the first reviewer was unsure whether to include/exclude were screened by a second reviewer, therefore in total, 14% of all titles were double screened at title and abstract.

### 5.4 Data extraction

Data extracted was conducted by a single reviewer and was consistency checked by a second reviewer. Information extracted includes:

- Reference (author, year, country)
- Study design
- Study aim
- Intervention details (including type of intervention/duration/delivery method/setting)
- Comparator intervention/ control
- Data collection methods and dates
- Outcomes
- Sample size
- Participants
- Anxiety and/or depression diagnosis and classification method
- Smoking abstinence outcomes
- Mental health
- Adverse events outcomes
- Observation/ notes

### 5.5 Study design classification

A study classification algorithm was not necessary for this rapid review.

### 5.6 Quality appraisal

The JBI critical appraisal checklists for quasi-experimental studies (updated 2024 version) and RCTs were used to assess the methodological quality of each included study. These checklists are not designed to assign an overall score to each study.

Quality assessment was undertaken in duplicate by two independent reviewers. Any discrepancies were discussed and resolved between reviewers. The quality assessment of individual studies can be seen in section 6.3.

### 5.7 Synthesis

Data was synthesised narratively to provide a collective interpretation of the evidence.

### 5.8 Assessment of body of evidence

To assess the body of evidence, findings for each category of interventions were grouped into smoking abstinence outcomes, smoking reduction outcomes, mental health outcomes, adverse events, and cost-effectiveness. The overall quality of studies reporting each outcome was considered in addition to the consistency of study findings in favour of the intervention, to assess whether the certainty of the evidence was considered very low, low, moderate or high.

## EVIDENCE

### 6.1 Search results and study selection

A total of 7,778 records were retrieved which were managed in Endnote 20. Following deduplication, 3,867 records remained. The search strategy for Medline is available in appendix 1. A total of 140 articles were screened at full text by two independent reviewers, and any conflicts were discussed and resolved within the team. A visual representation of the flow of studies throughout the review can be found in Figure 1.

**Figure 1.**
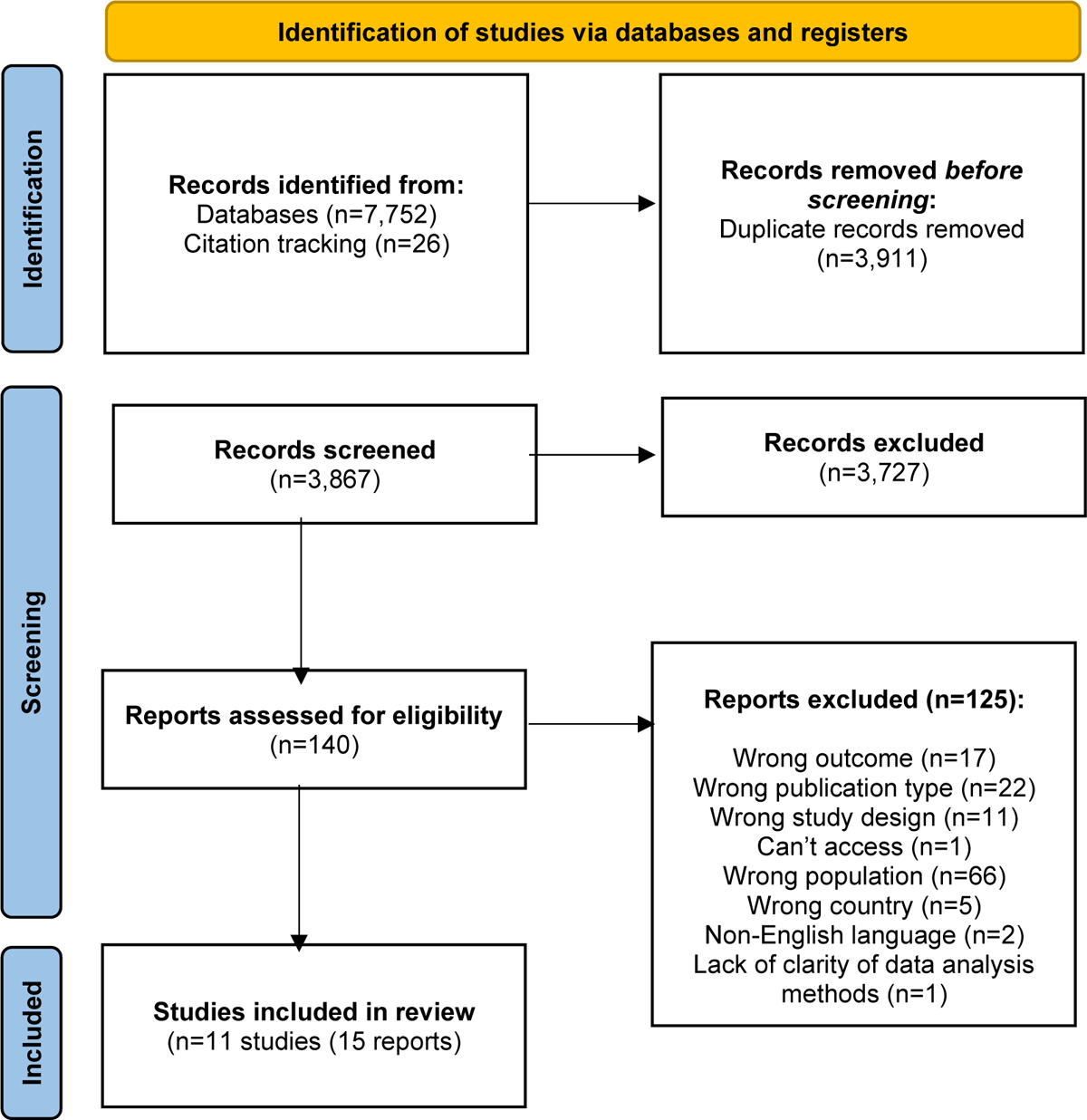
PRISMA flow diagram

Eleven studies reported in 15 publications (reports) were include in the rapid review. Ten of the 11 studies were RCTs, and one was a quasi-experimental uncontrolled before and after pilot study.

### 6.2 Data extraction

**Table 5.**
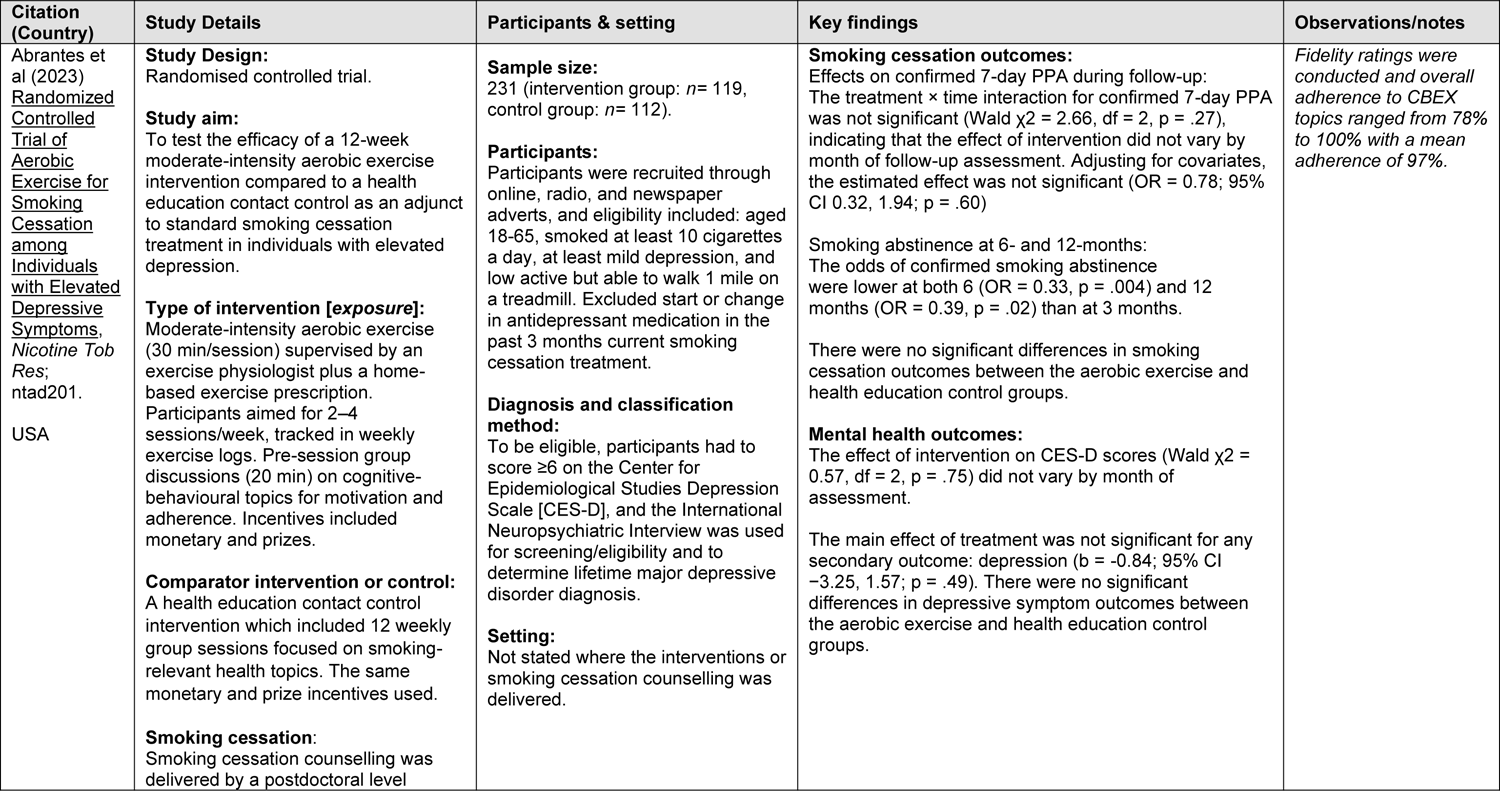
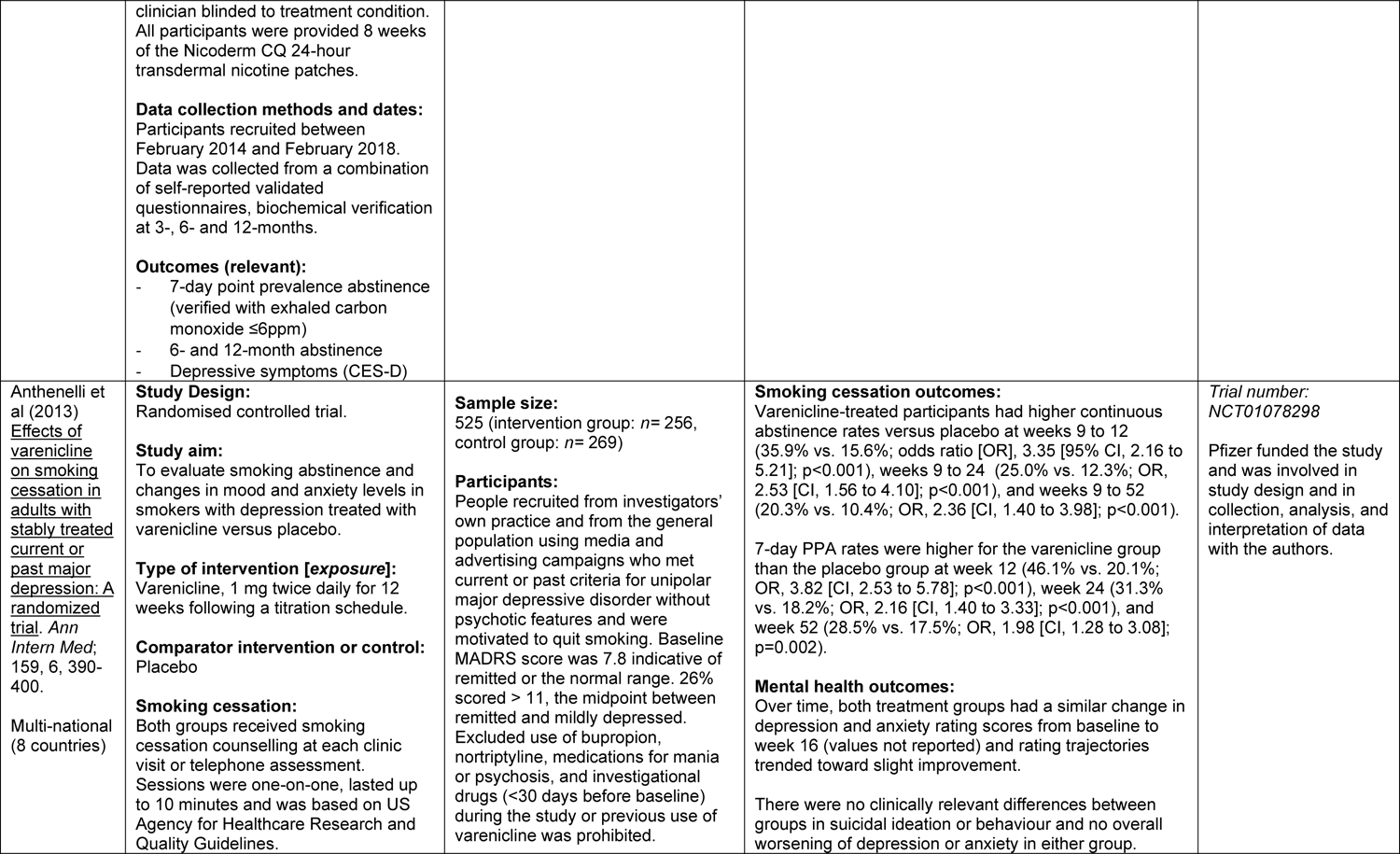
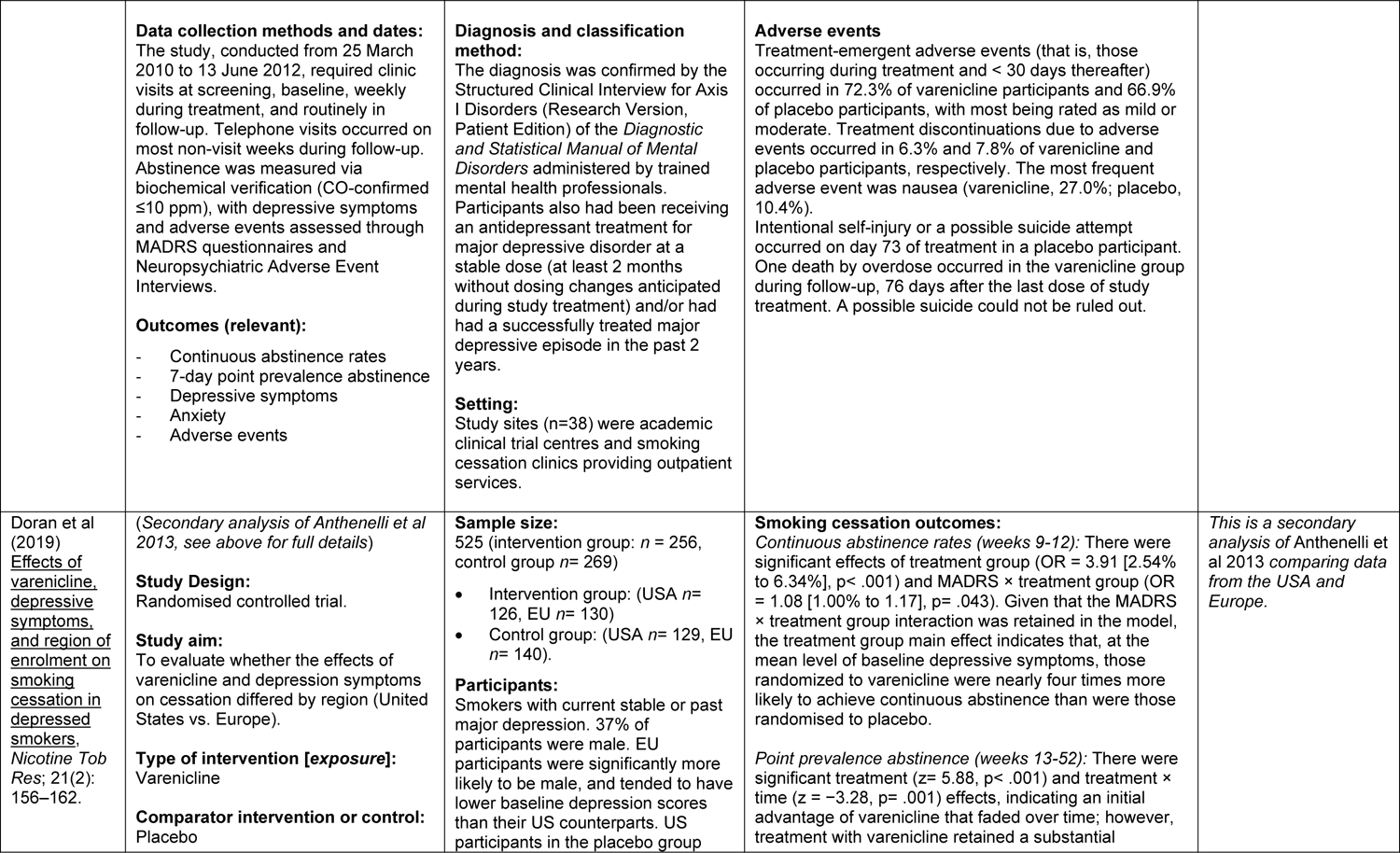
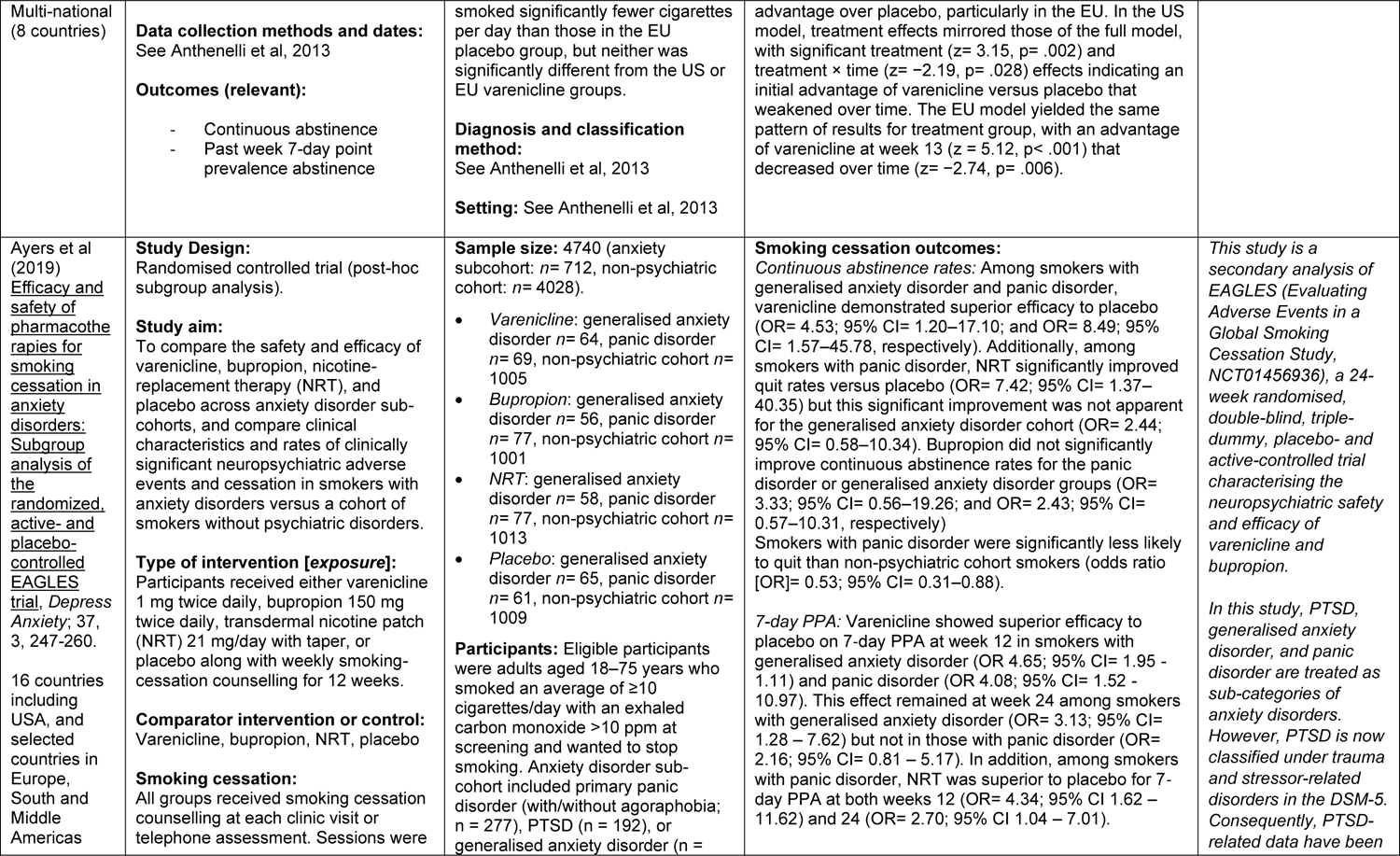
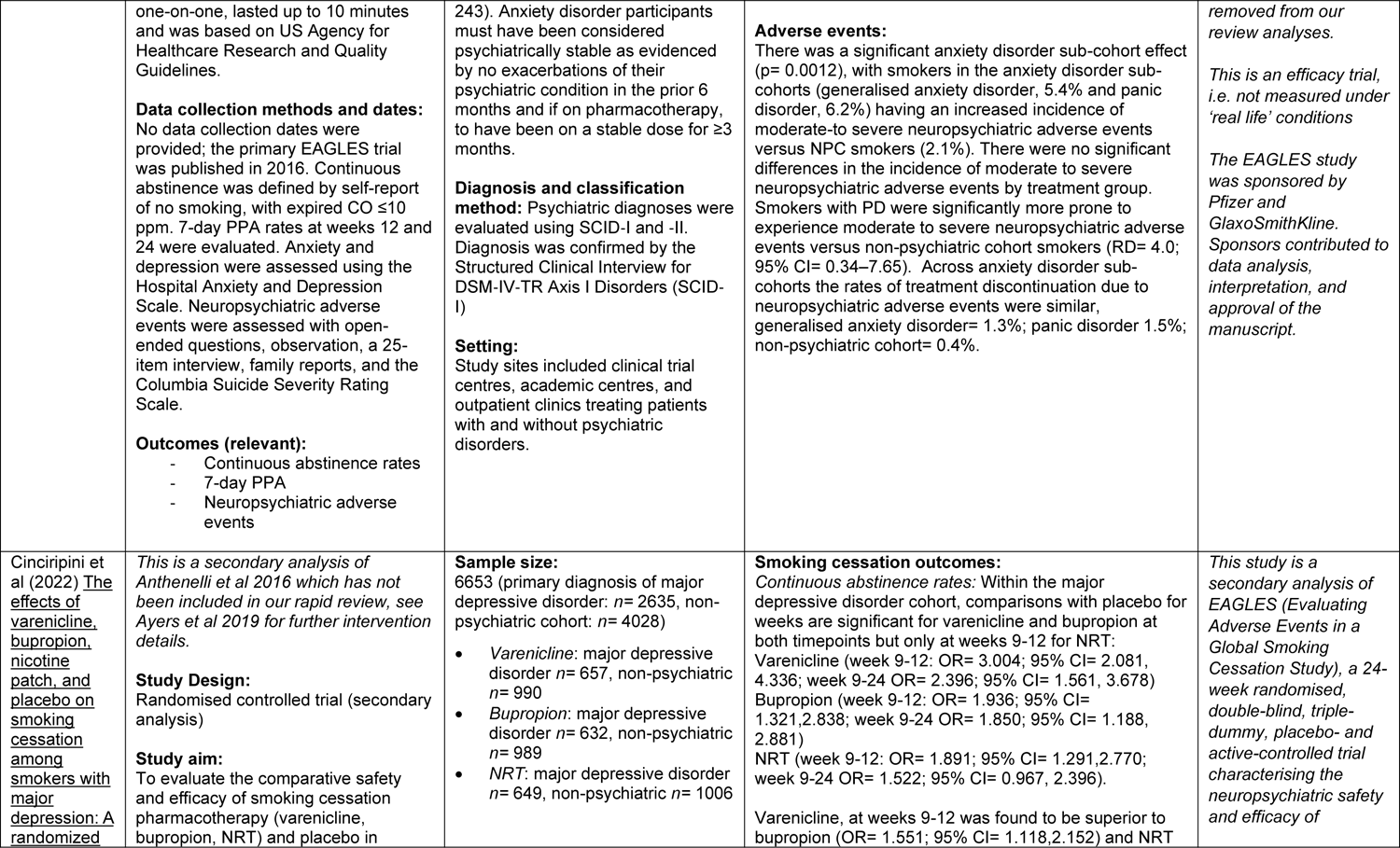
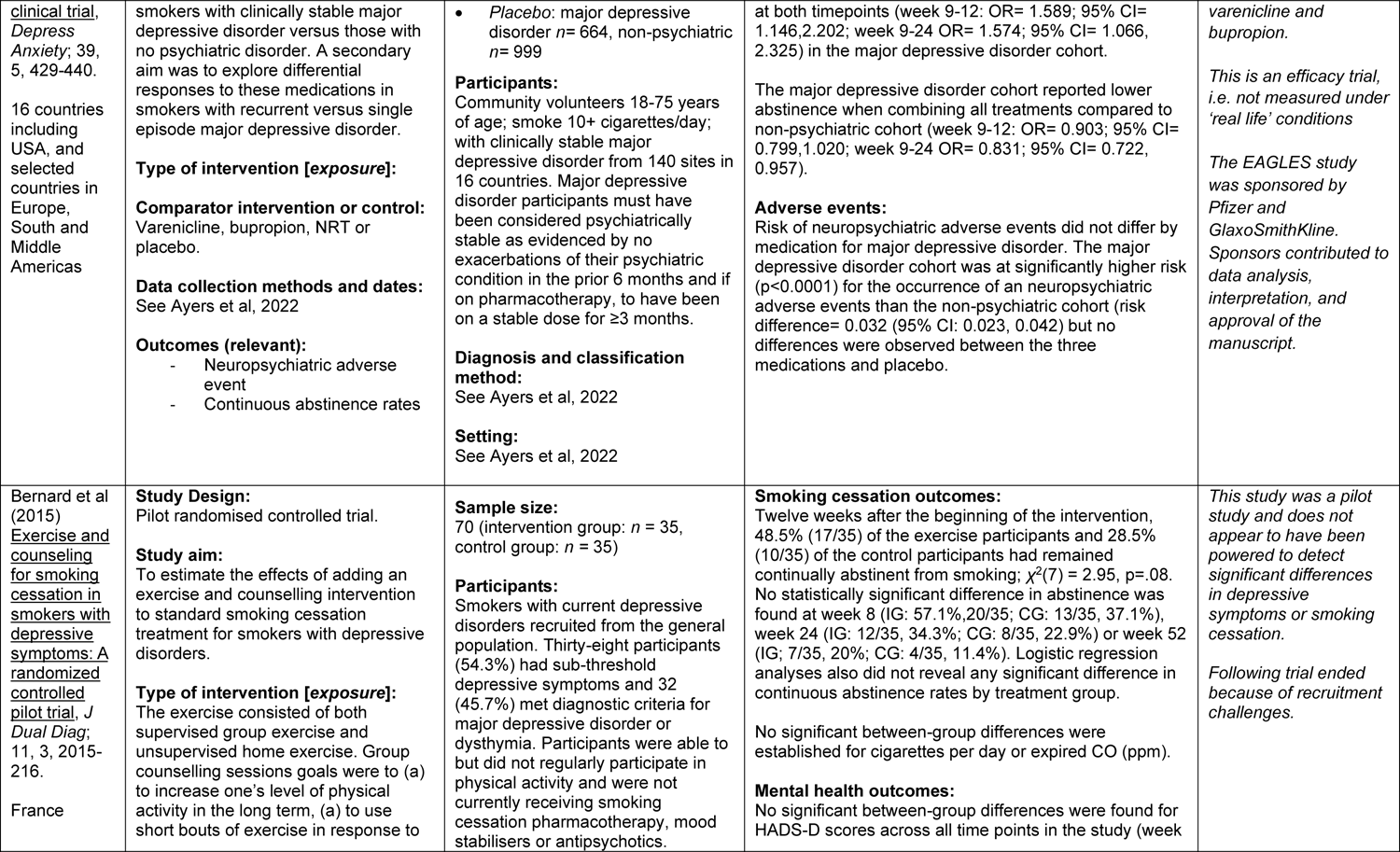
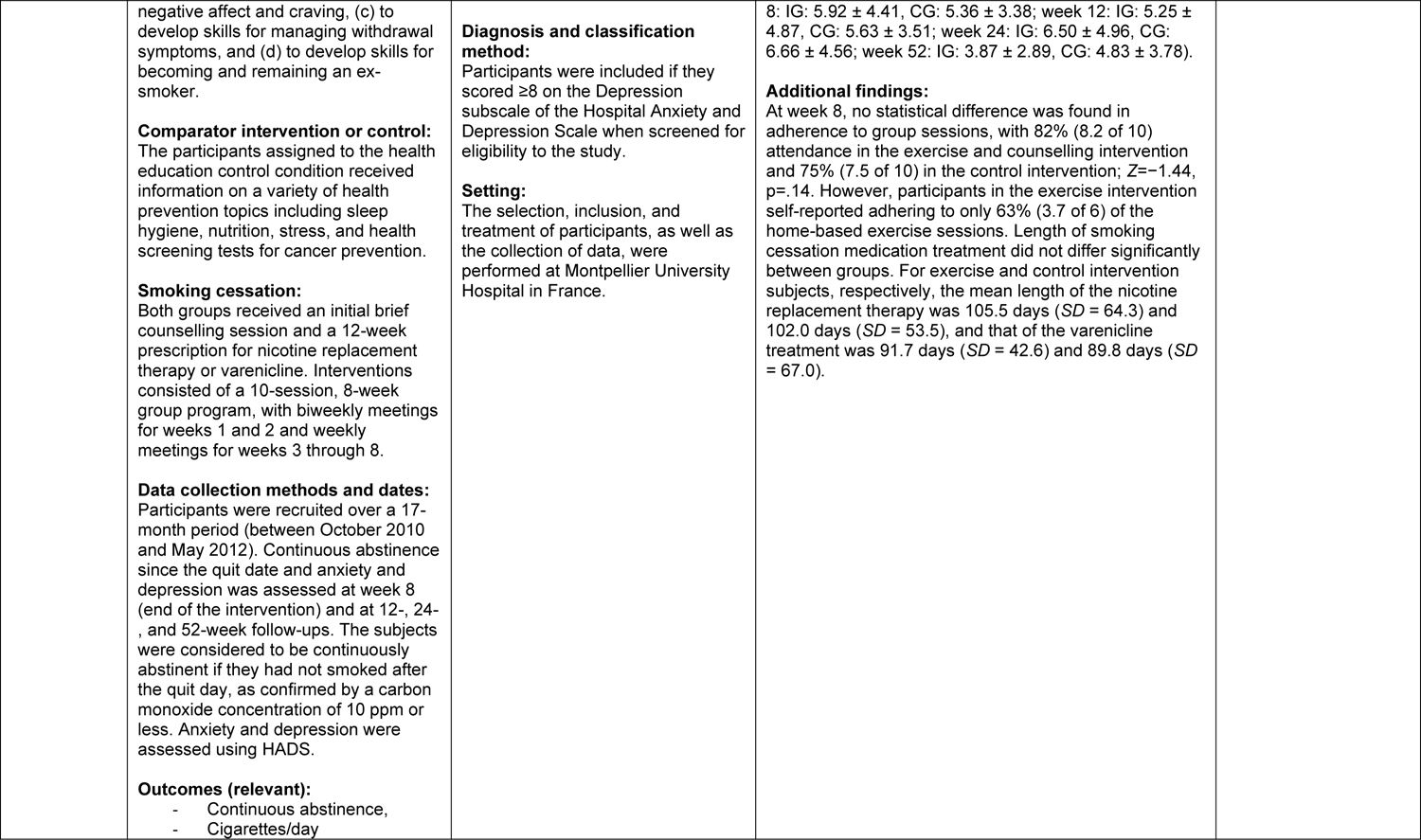
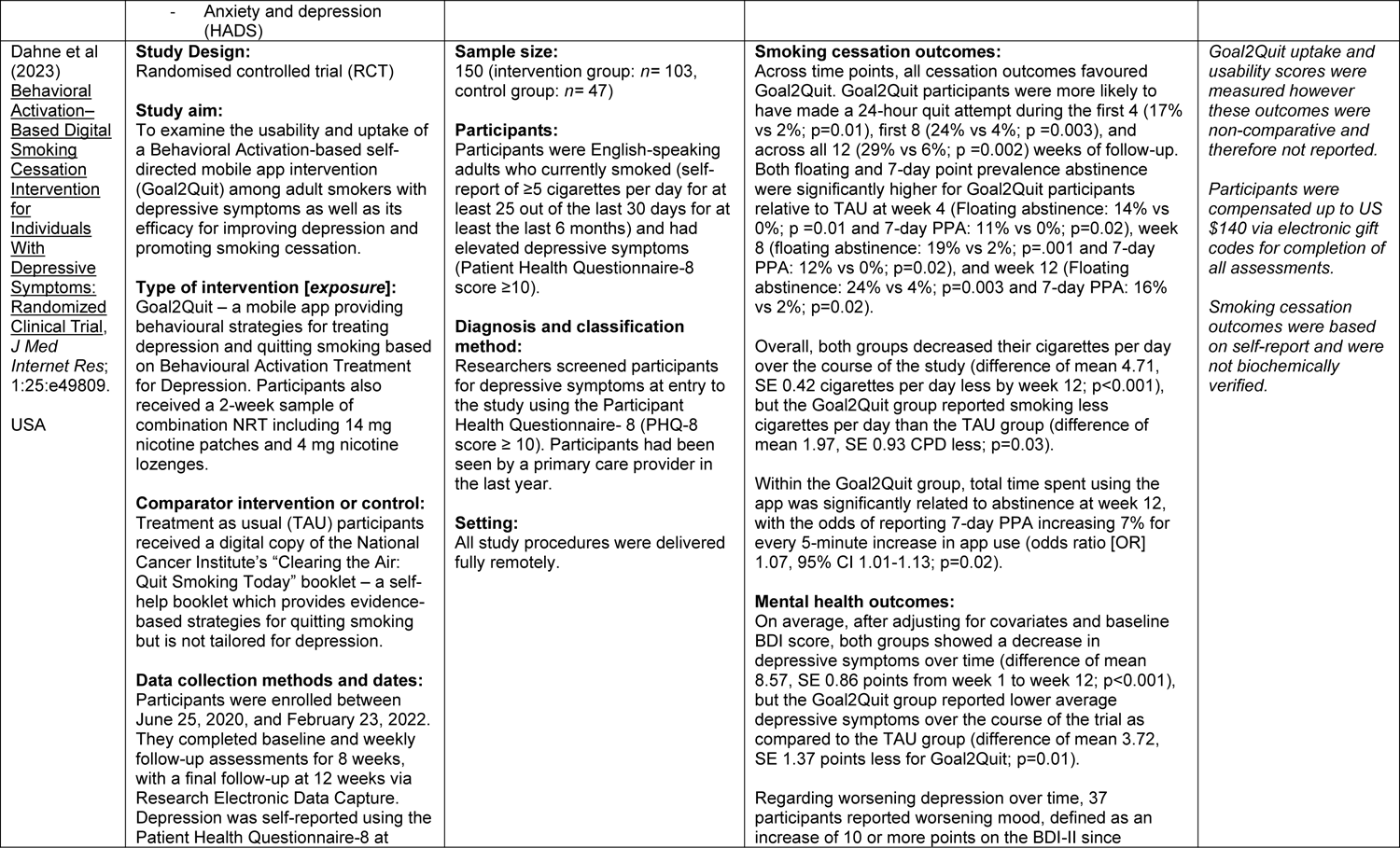
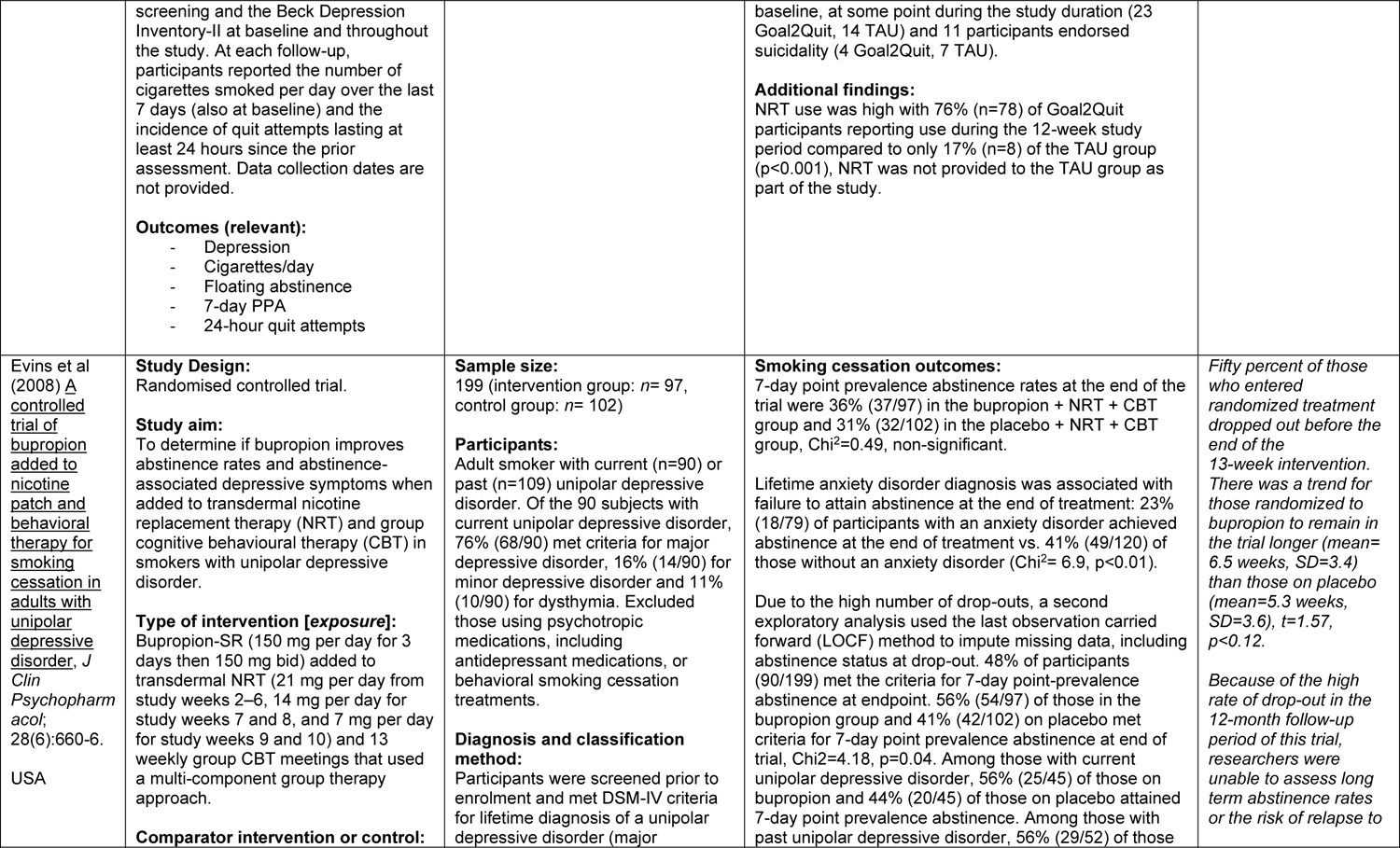
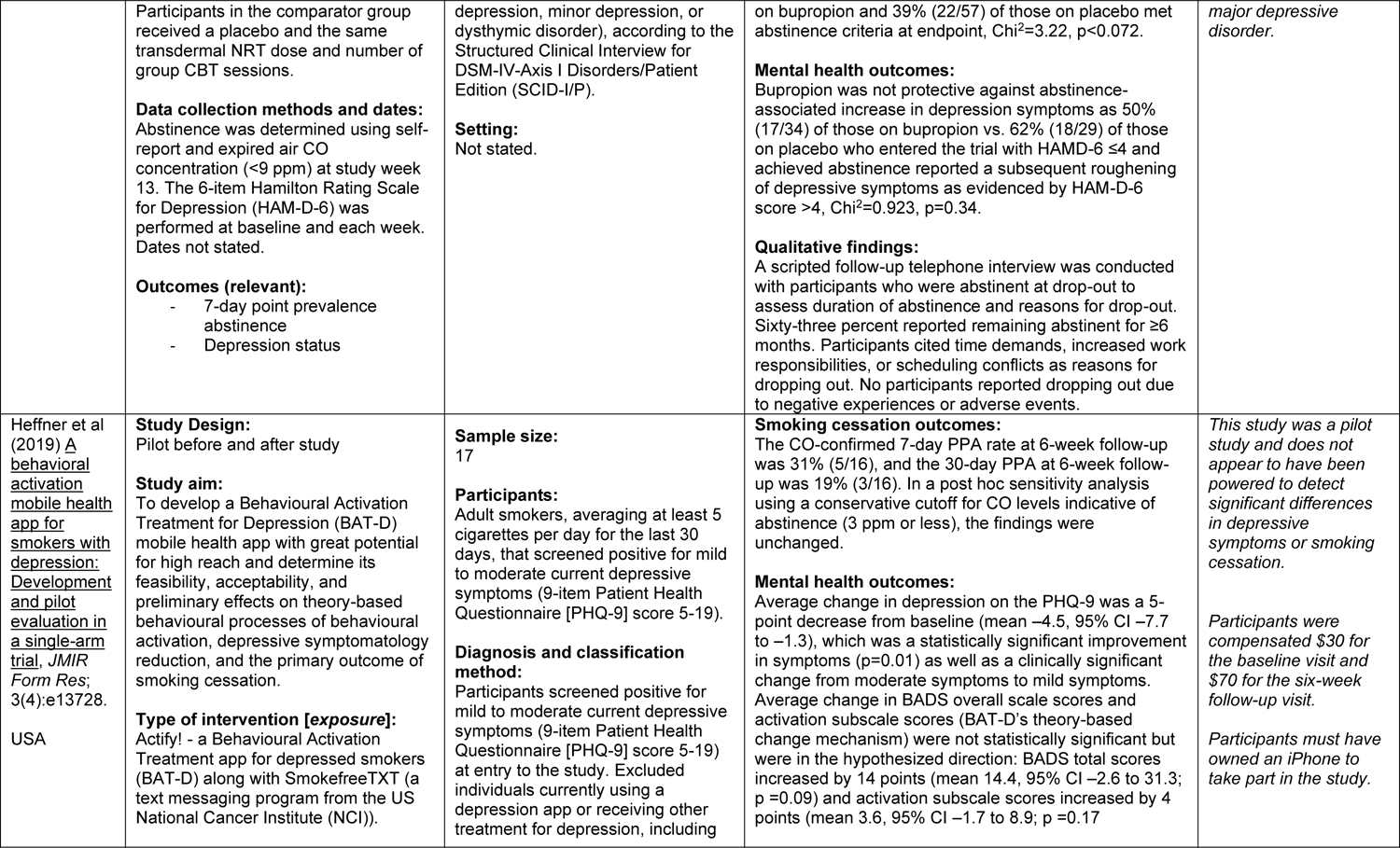
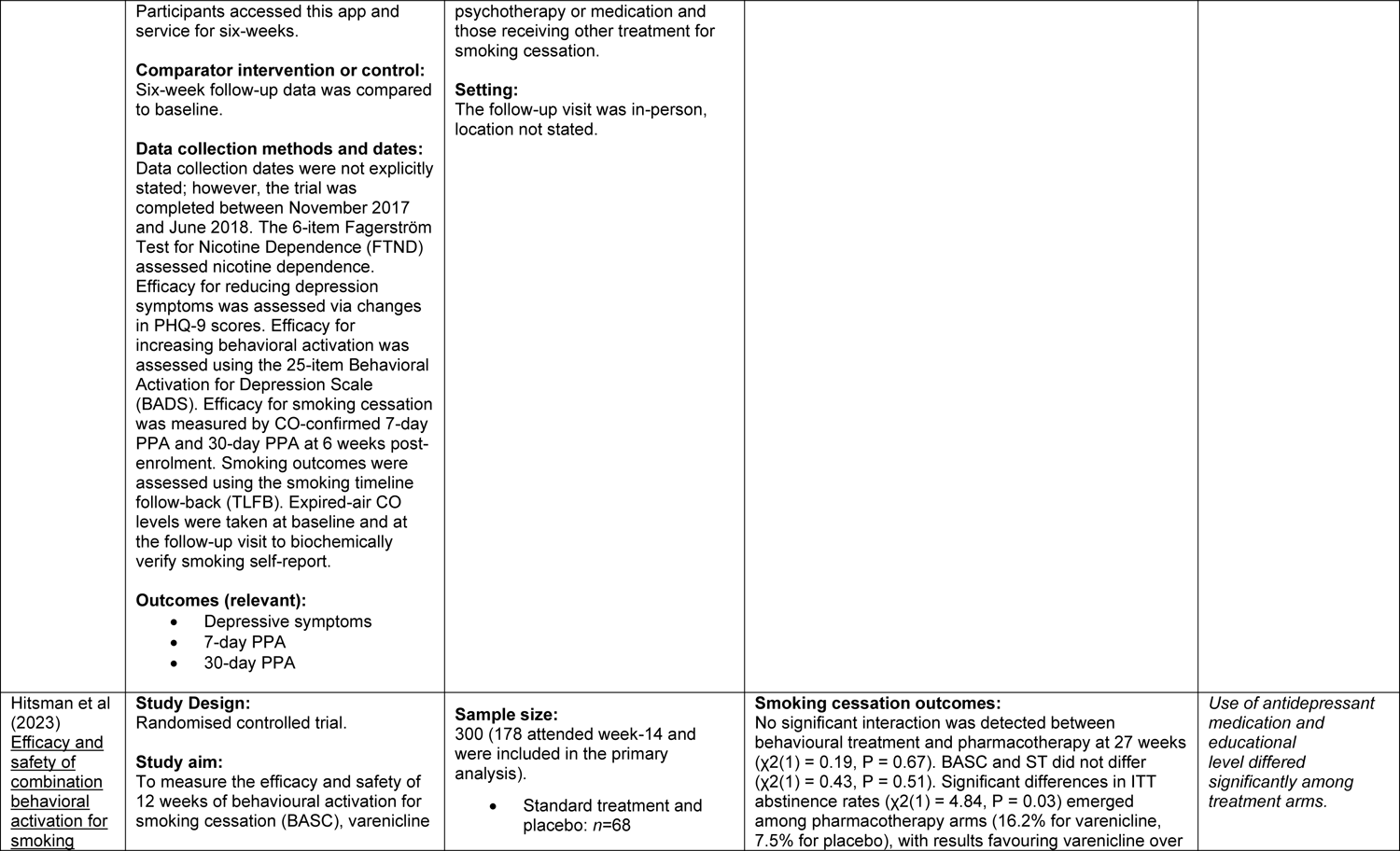
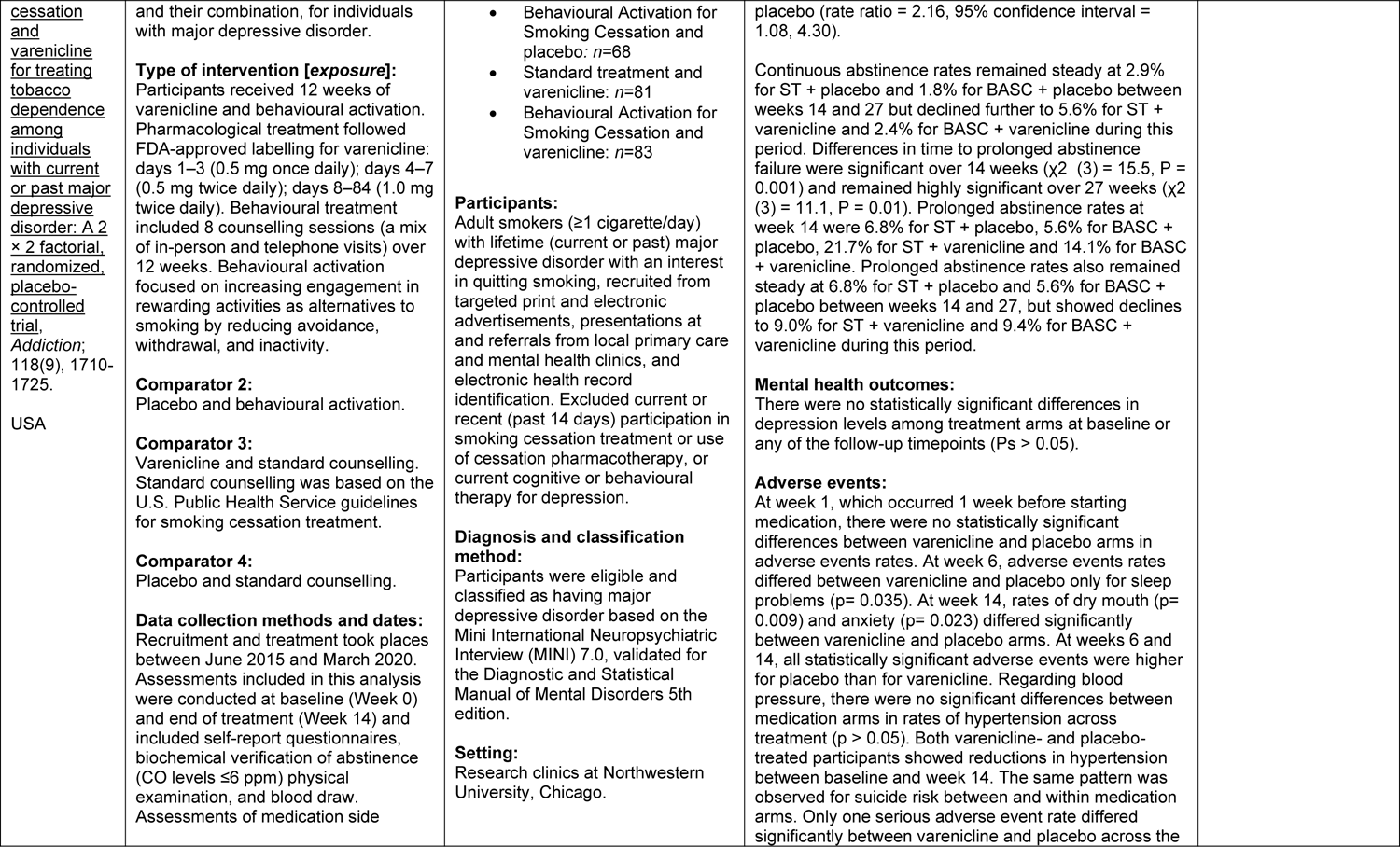
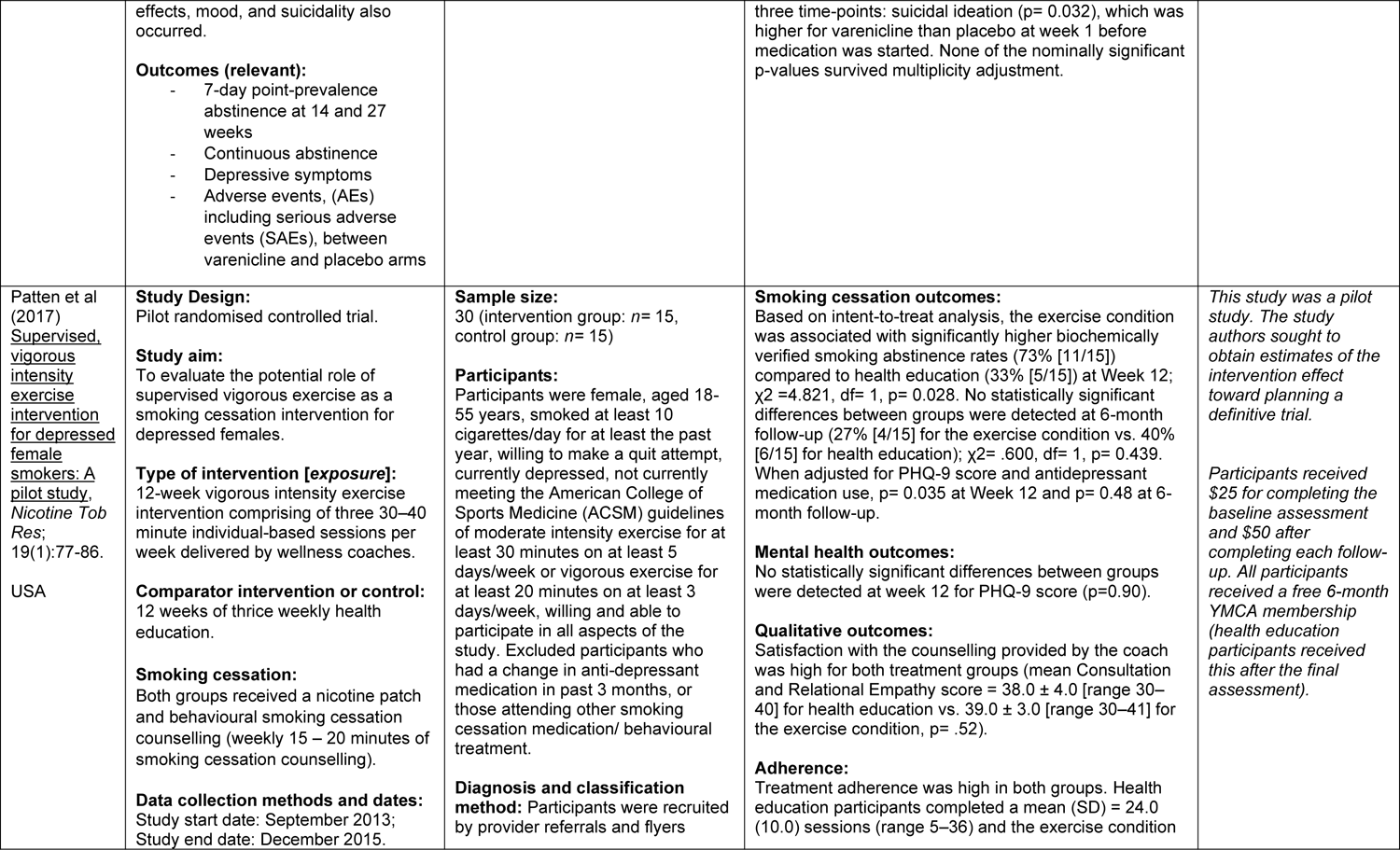
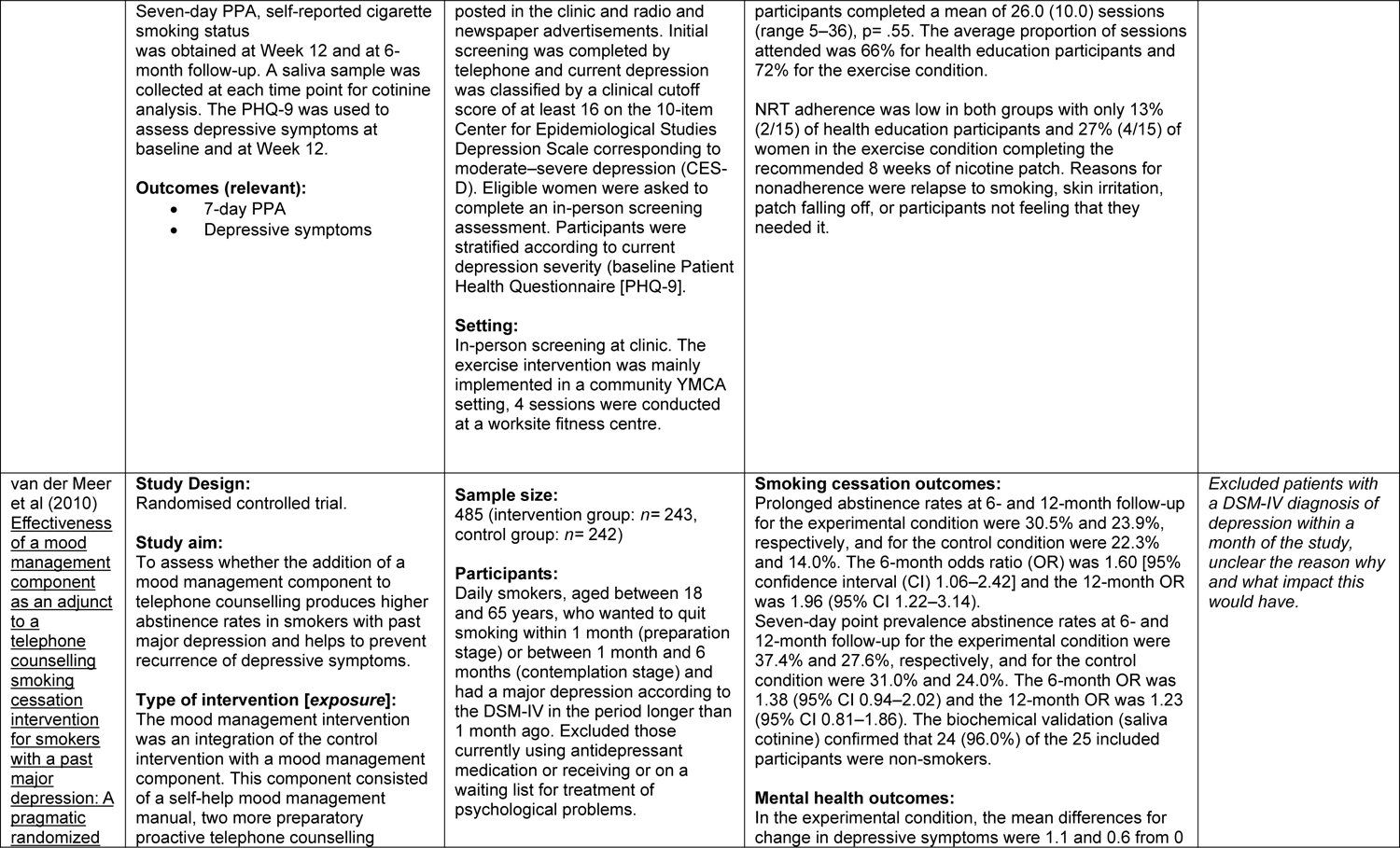
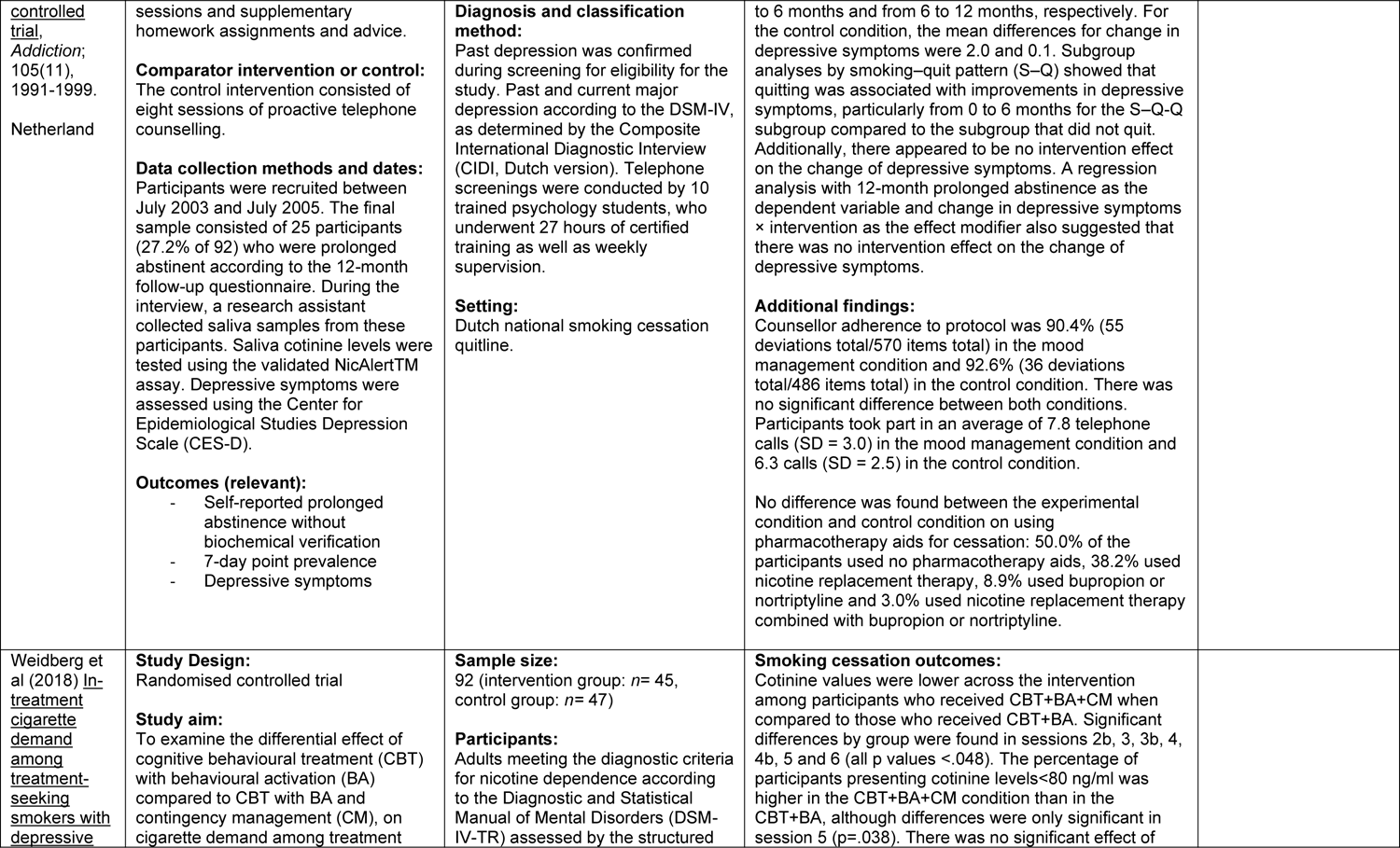
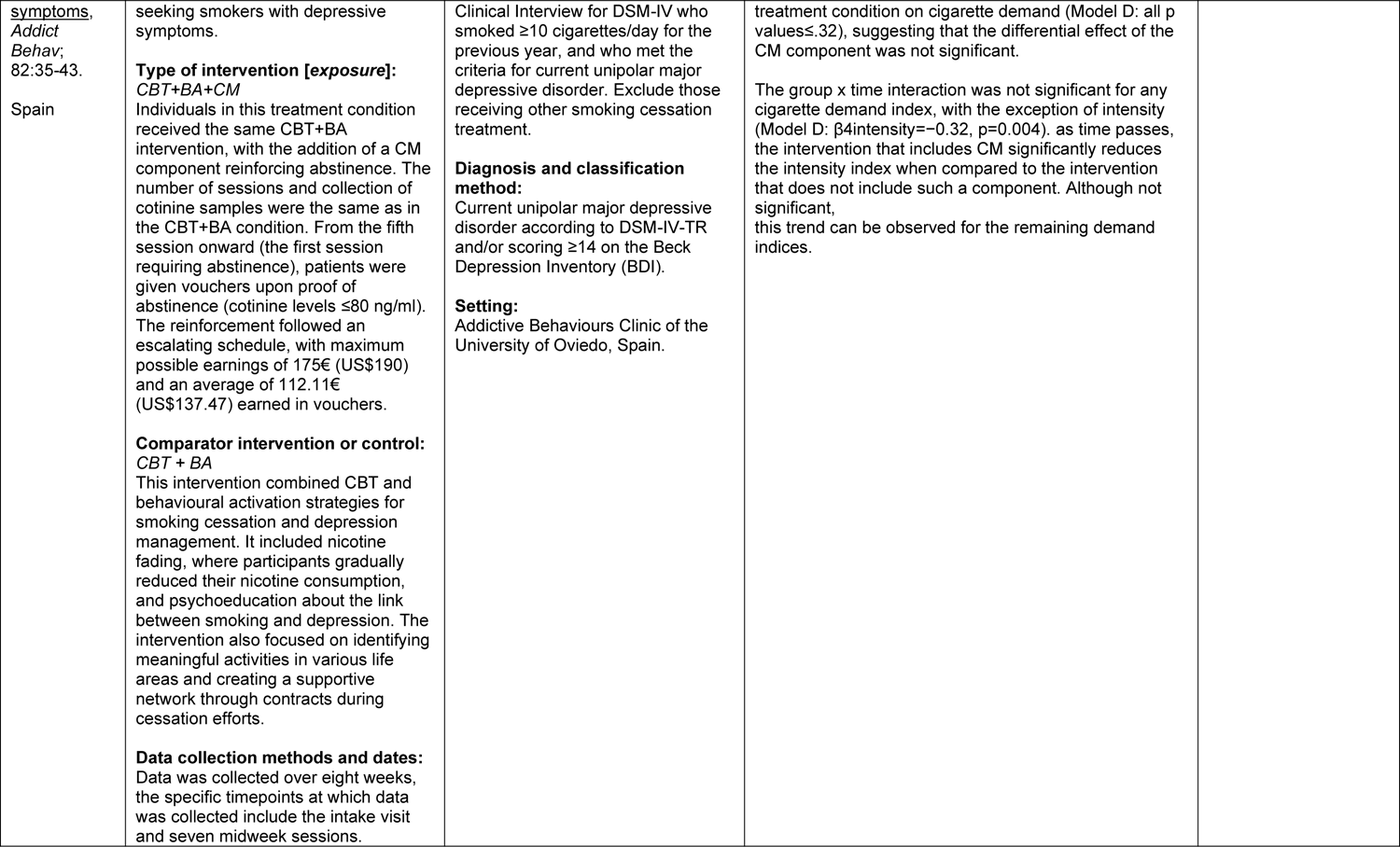
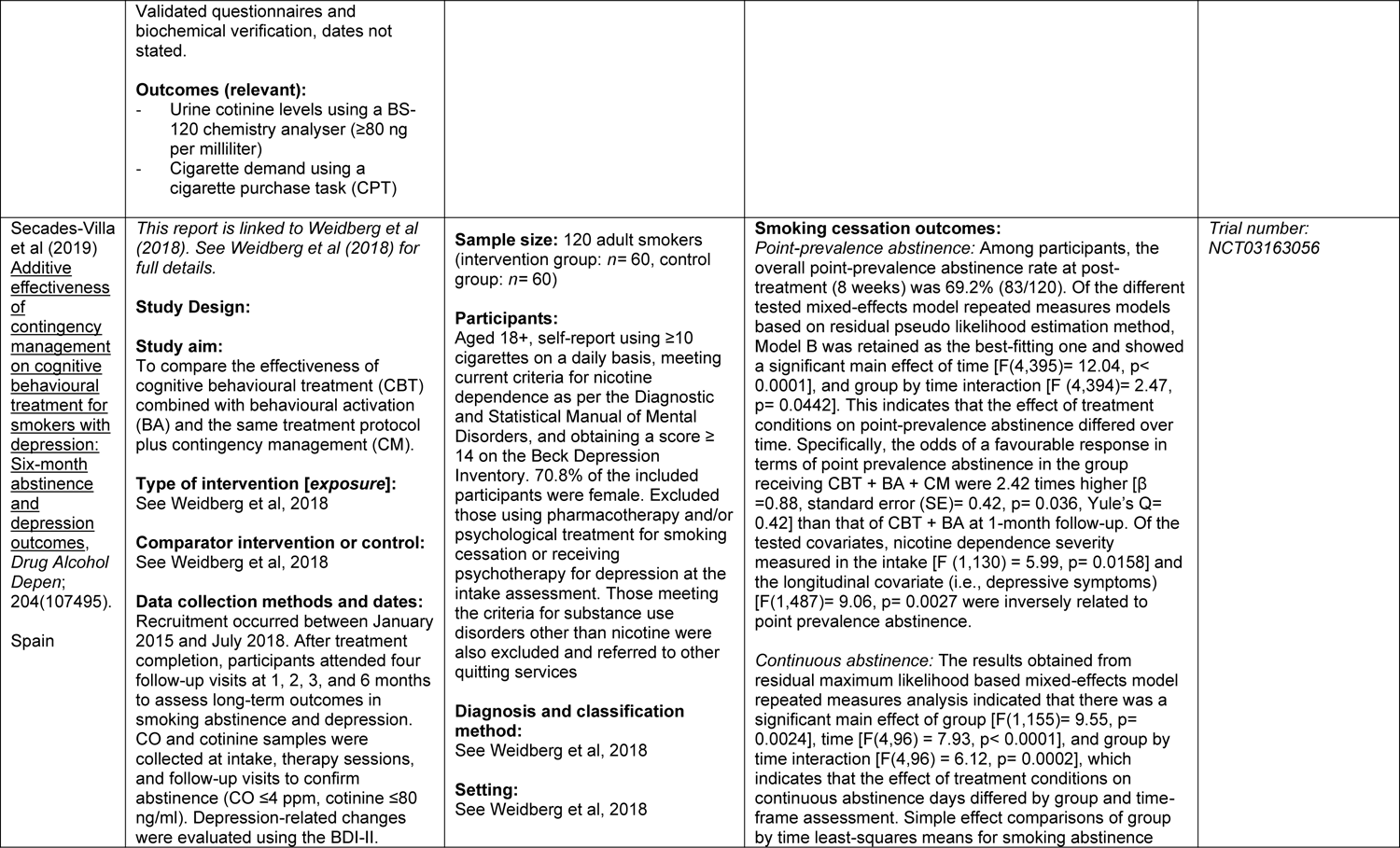
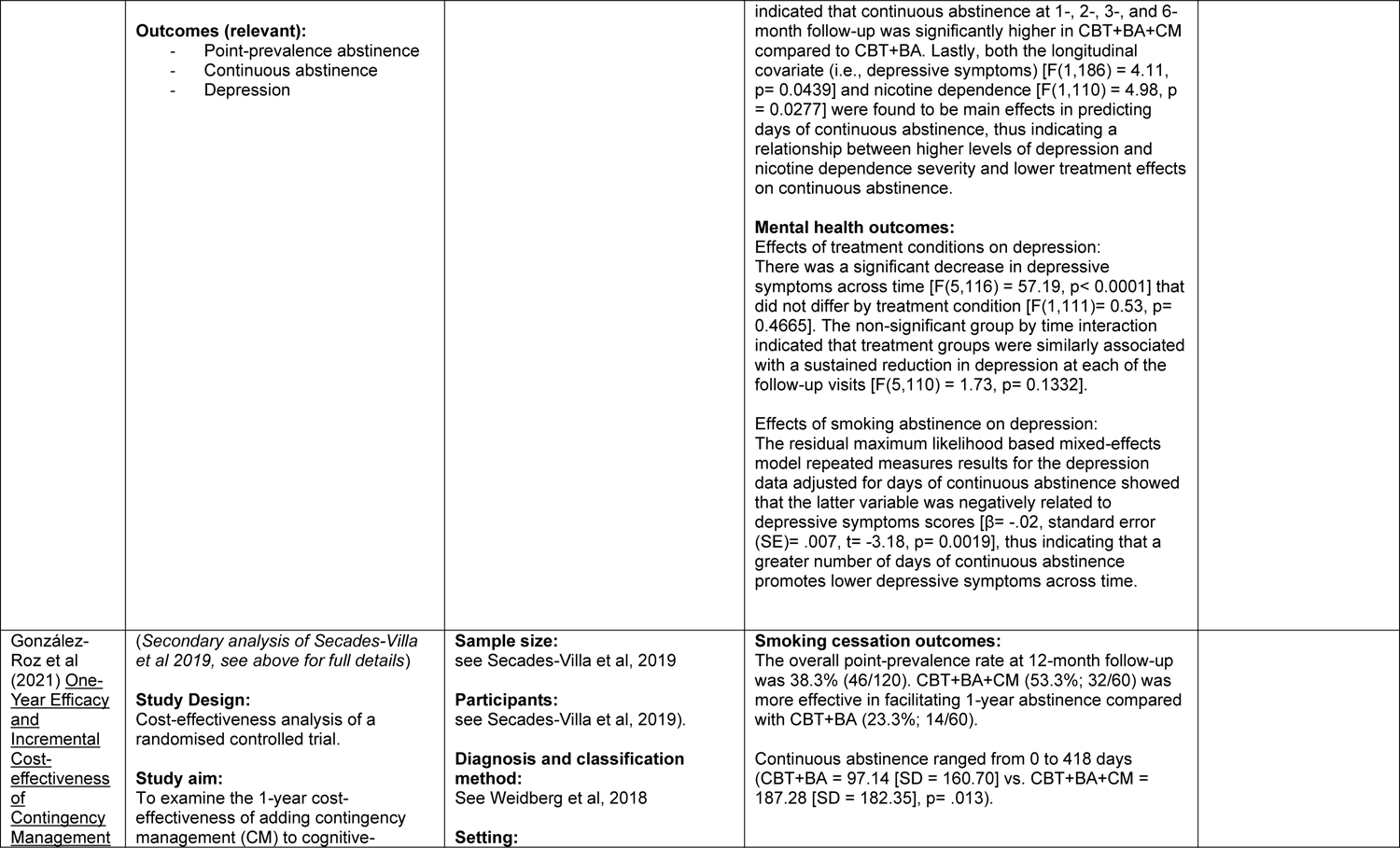
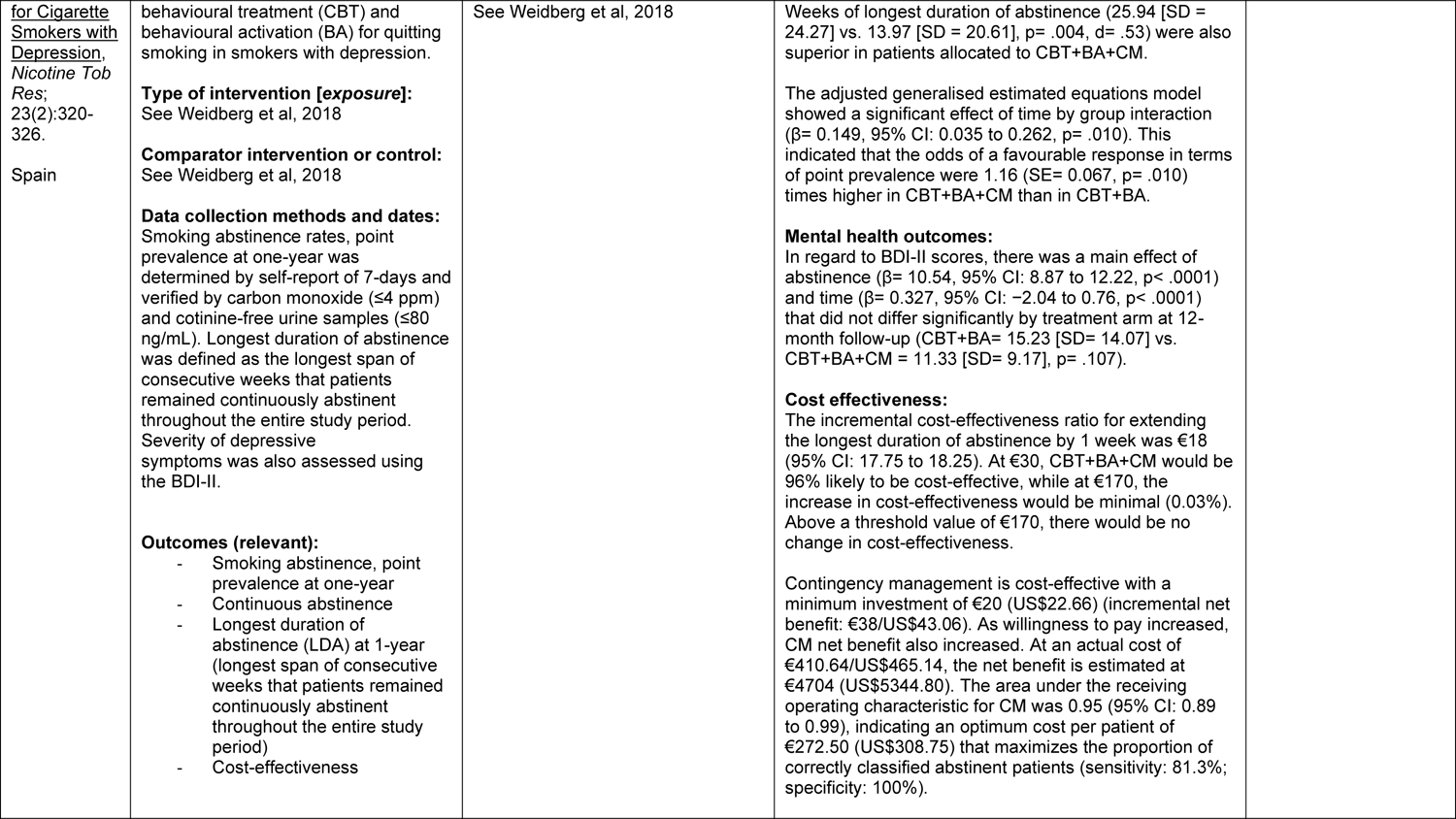
Summary of included studies.

### 6.3 Quality appraisal

**Table 6.**
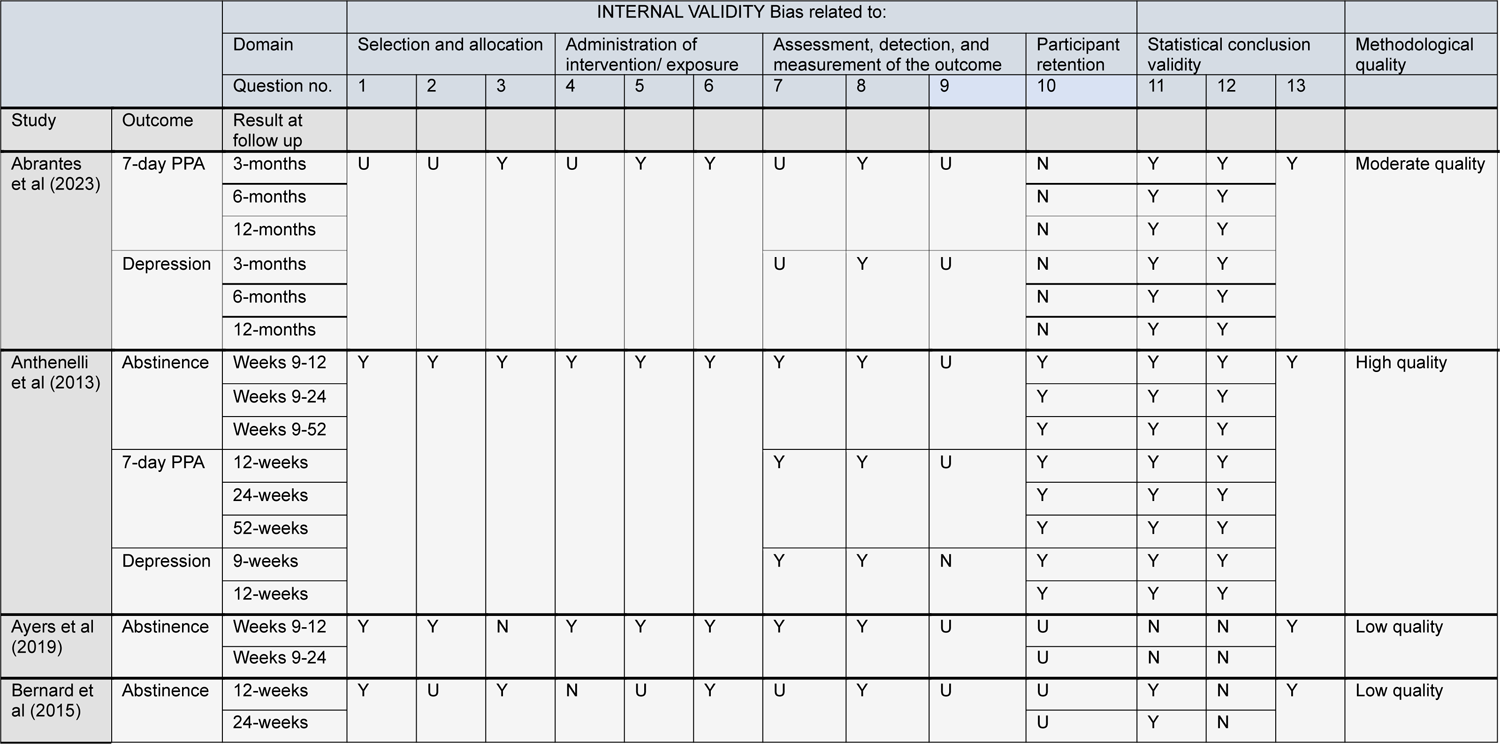
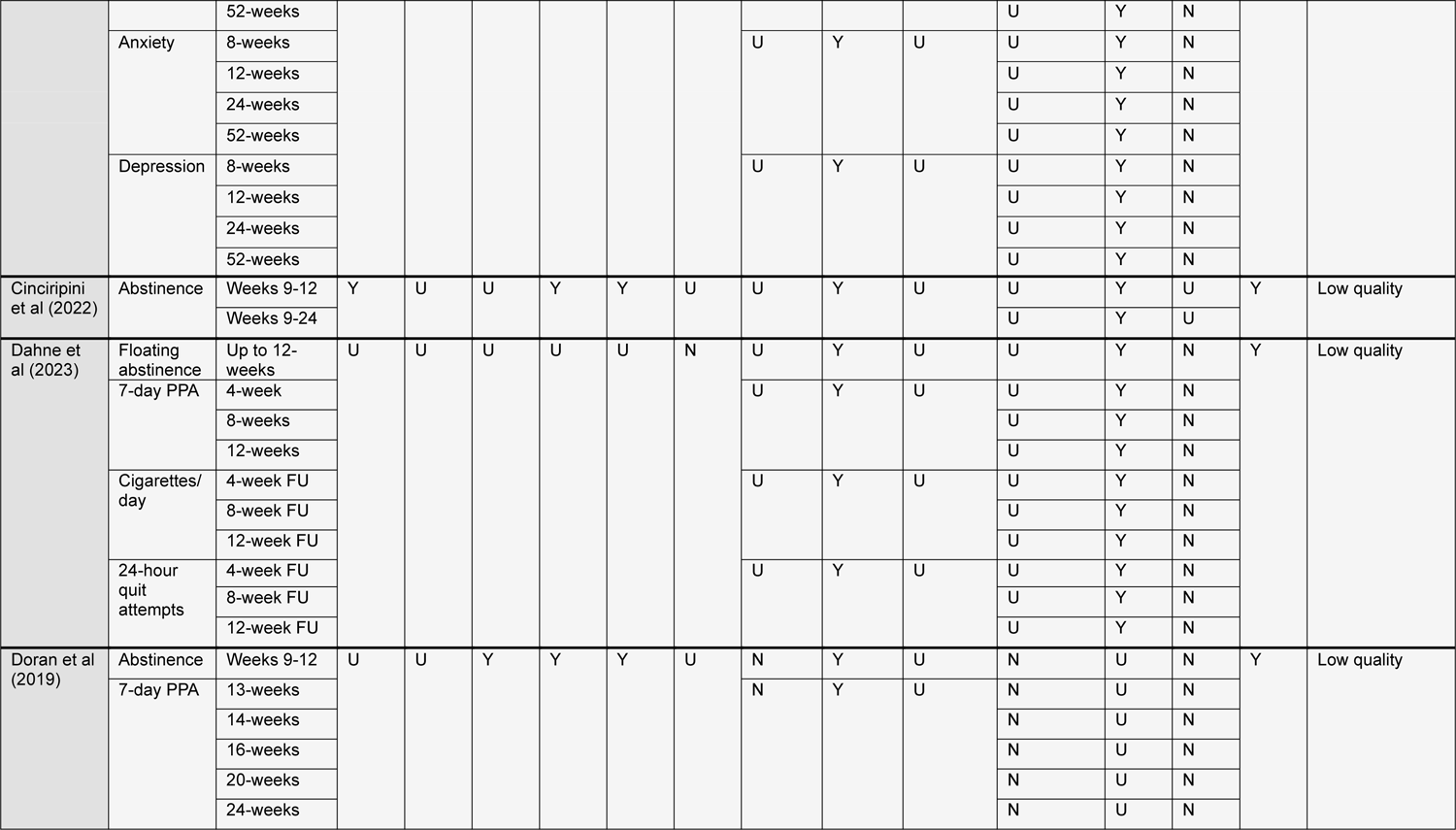
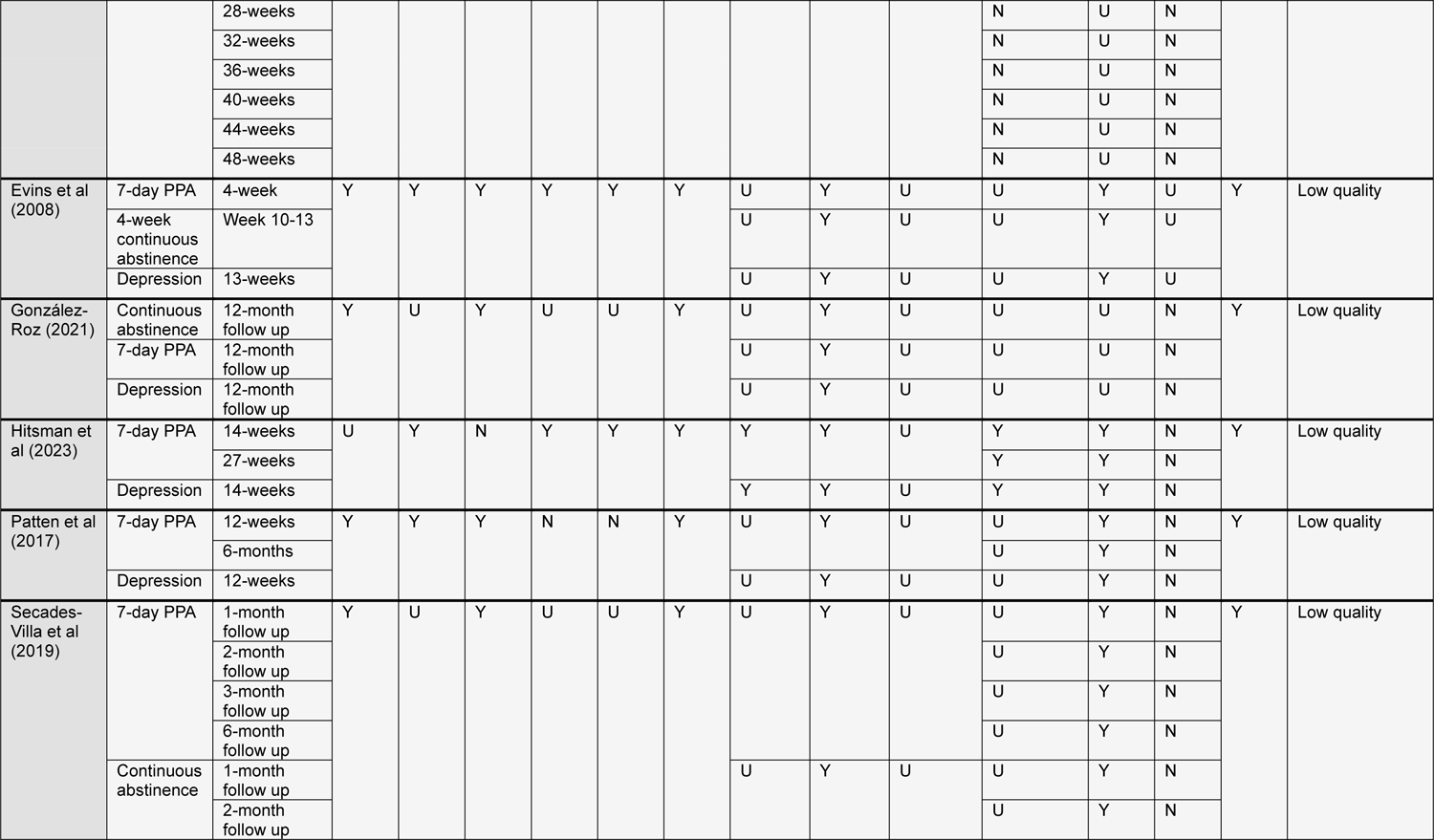
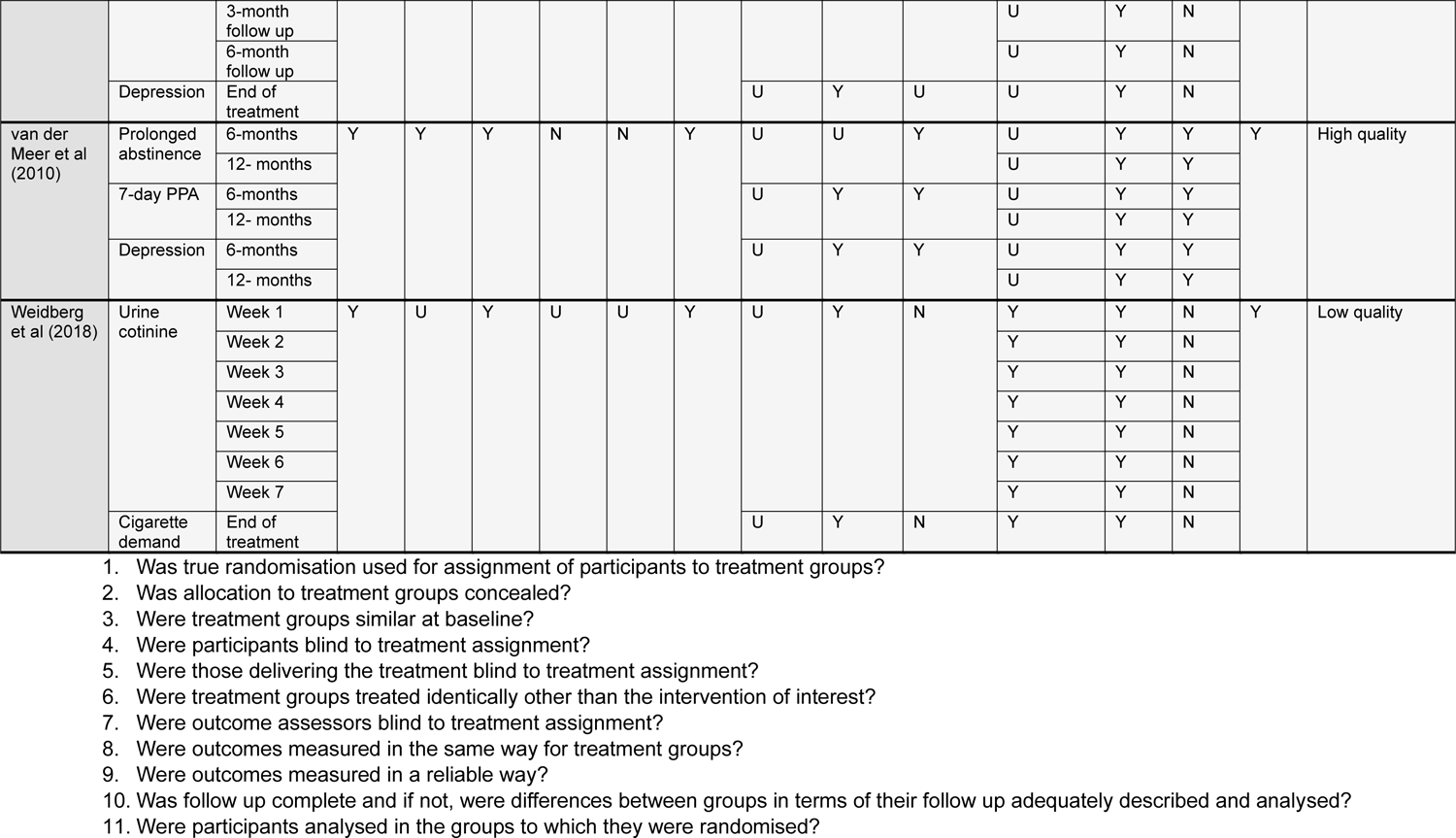
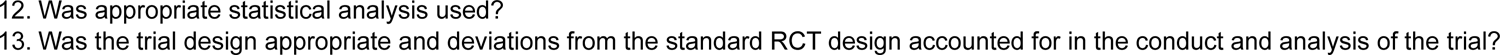
Risk of bias of randomised controlled trials.

**Table 7.**
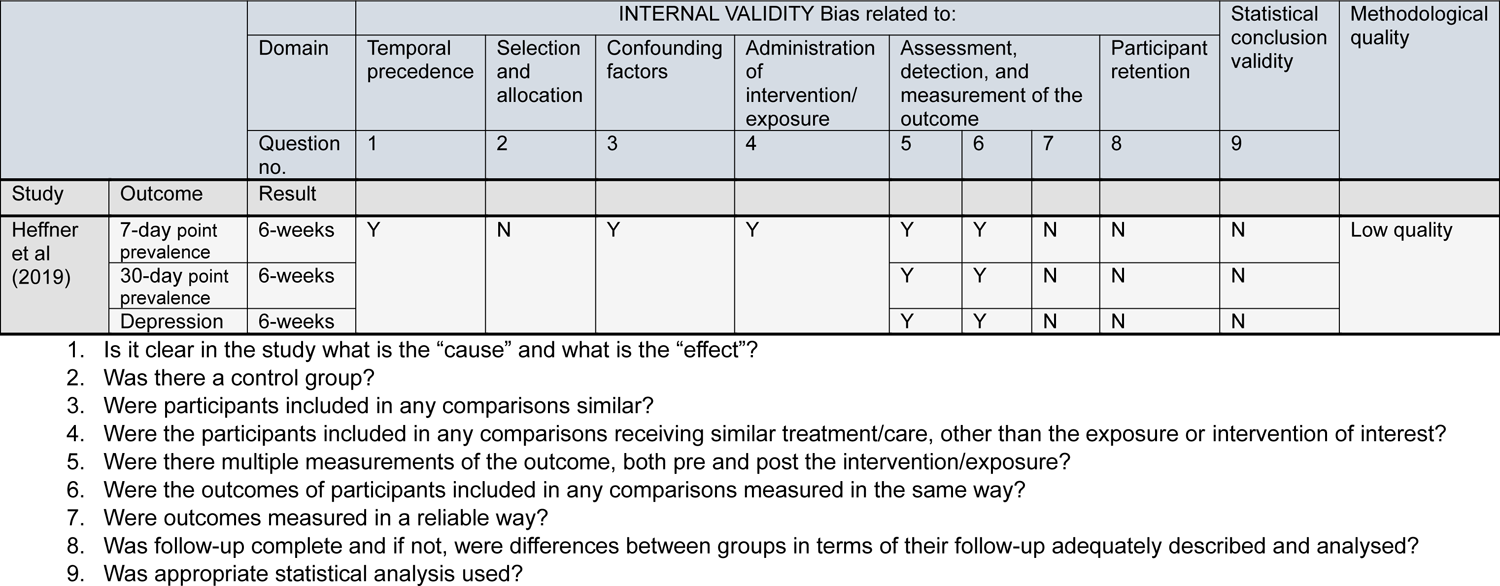
Risk of bias of quasi-experimental studies.

### 6.4 Information available on request

The following are available on request: protocol; search strategies.

## 7.2 Acknowledgements

The Public Health Wales team would like to thank Chris Emmerson, Chris Roberts, Claire Pritchard, Josie Jackson, Elizabeth Doe, Liz Newbury-Davies, Lorna Bennett, Nathan Davies, Olivia Gallen, Orla Thomas, Rachael Thatcher, Stephanie Barnhouse and Wallis Jones, for their time, expertise and contributions during stakeholder meetings in guiding the focus of the review and interpretation of findings.

## APPENDIX

### APPENDIX 1

Ovid MEDLINE(R) ALL <1946 to March 20, 2024>

1. (anxiety or anxious or depression or depressed or depressive).ti,ab. 690559
2. exp Anxiety/ or Anxiety Disorders/ or Depression/ or Anxiety, Separation/ or Panic Disorder/ or exp Phobic Disorders/ 277678
3. 1 or 2 749926
4. Smoking Cessation/ or “Tobacco Use Cessation”/ or “Tobacco Use Cessation Devices”/ 34902
5. (”smoking cessation” or “tobacco cessation” or (tobacco adj2 abstain*) or (smok* adj2 abstain*) or (cigar* adj2 abstain*) or (tobacco adj2 abstinence) or (smok* adj2 abstinence) or (cigar* adj2 abstinence) or (stop* adj2 tobacco) or (stop* adj2 smok*) or (stop* adj2 cigar*) or (quit* adj2 tobacco) or (quit* adj2 smok*) or (quit* adj2 cigar*) or (ceas* adj3 tobacco) or (ceas* adj3 smok*) or (ceas* adj3 cigar*) or (”give up” adj2 tobacco) or (”give up” adj2 smok*) or (”give up” adj2 cigar*)).ti,ab. 43148
6. smoking reduction/ 116
7. (”smoking reduc*” or “tobacco reduc*” or (tobacco adj3 “cut down”) or (smoking adj3 “cut down”) or (cigar* adj3 “cut down”)).ti,ab. 1540
8. 4 or 5 or 6 or 7 55485
9. 3 and 8 3192
10. Clinical Study/ or exp Clinical Trial/ or Clinical Trial Protocol/ or Observational Study/ or Comparative Study/ 2848018
11. (treatment* or approach* or strateg* or intervention* or trial*).ti,ab. 9710019
12. 10 or 11 11404694
13. 9 and 12 2317
14. limit 13 to (english language and yr=”2007-Current”) 1741
15. (case reports or comment or editorial or letter).pt. 4394954
16. exp animals/ not humans/ 5205065
17. 15 or 16 9488032
18. 14 not 17 1675

### APPENDIX 2

A reference list of smoking cessation studies comparing people with mental health conditions to the general population. These studies were identified by our search and deemed ineligible for this rapid review but may contribute to a broader understanding of the impact of these interventions in this population.

#### Depression populations

##### Pharmacological and psychological interventions

McClure, J. B., Swan, G. E., Jack, L., Catz, S. L., Zbikowski, S. M., McAfee, T. A., … & Javitz, H. (2009). Mood, side-effects and smoking outcomes among persons with and without probable lifetime depression taking varenicline. *Journal of general internal medicine*, *24*, 563-569. Available at: https://link.springer.com/article/10.1007/s11606-009-0926-8 *Comparing those with probable lifetime major depression vs no apparent major depression history*.

##### Pharmacological interventions

Minami, H., Kahler, C. W., Bloom, E. L., Strong, D. R., Abrantes, A. M., Zywiak, W. H., … & Brown, R. A. (2015). Effects of depression history and sex on the efficacy of sequential versus standard fluoxetine for smoking cessation in elevated depressive symptom smokers. *Addictive Disorders & Their Treatment*, *14*(1), 29-39. Available at: https://journals.lww.com/addictiondisorders/fulltext/2015/03000/Effects_of_Depression_History_and_Sex_on_the.3.aspx *Comparing those with no past depressive episode vs recurrent depressive episodes vs single past depressive episode*.

Spring, B., Doran, N., Pagoto, S., McChargue, D., Cook, J. W., Bailey, K., … & Hedeker, D. (2007). Fluoxetine, smoking, and history of major depression: A randomized controlled trial. *Journal of consulting and clinical psychology*, *75*(1), 85. Available at: https://core.ac.uk/reader/17225910 *Comparing those with a history of depression vs without history of depression*.

Tidey, J. W., Pacek, L. R., Koopmeiners, J. S., Vandrey, R., Nardone, N., Drobes, D. J., … & Donny, E. C. (2016). Effects of 6-week use of reduced-nicotine content cigarettes in smokers with and without elevated depressive symptoms. *Nicotine & Tobacco Research*, *19*(1), 59-67. Available at: https://academic.oup.com/ntr/article/19/1/59/2631697 *Comparing those with vs without elevated depressive symptoms*.

Zawertailo, L., Voci, S., & Selby, P. (2015). Depression status as a predictor of quit success in a real-world effectiveness study of nicotine replacement therapy. *Psychiatry research*, *226*(1), 120-127. Available at: https://www.clinicalkey.com/#!/content/playContent/1-s2.0-S0165178114010208?returnurl=null&referrer=null *Participants were divided into four mutually exclusive groups: current/recent depression, recurrent depression, past depression and no depression history*.

Zhang, H., Gilbert, E., Hussain, S., Veldhuizen, S., Le Foll, B., Selby, P., & Zawertailo, L. (2023). Effectiveness of bupropion and varenicline for smokers with baseline depressive symptoms. *Nicotine and Tobacco Research*, *25*(5), 937-944. Available at: https://academic.oup.com/ntr/article/25/5/937/6911424 *Compares those with depressive symptoms vs those without*.

### Mixed mental health populations

#### Pharmacological and psychological interventions

Carroll, A. J., Mathew, A. R., Leone, F. T., Wileyto, E. P., Miele, A., Schnoll, R. A., & Hitsman, B. (2020). Extended nicotine patch treatment among smokers with and without comorbid psychopathology. *Nicotine and Tobacco Research*, *22*(1), 24-31. Available at: https://academic.oup.com/ntr/article/22/1/24/5095857 *Compares those with vs without a psychiatric condition*.

#### Psychological and exercise interventions

Garey, L., Rogers, A. H., Manning, K., Smit, T., Derrick, J. L., Viana, A. G., … & Zvolensky, M. J. (2020). Effects of smoking cessation treatment attendance on abstinence: The moderating role of psychologically based behavioral health conditions. *Journal of substance abuse treatment*, *109*, 1-7. Available at: https://www.ncbi.nlm.nih.gov/pmc/articles/PMC6927534/pdf/nihms-1542480.pdf *Compares those with vs without psychologically based behavioural health conditions*.

#### Media campaigns

Prochaska, J. J., Gates, E. F., Davis, K. C., Gutierrez, K., Prutzman, Y., & Rodes, R. (2019). The 2016 tips from former smokers® campaign: associations with quit intentions and quit attempts among smokers with and without mental health conditions. *Nicotine and Tobacco Research*, *21*(5), 576-583. Available at: https://academic.oup.com/ntr/article/21/5/576/5212283?login=true *Compares people living with mental health conditions vs people without mental health conditions*.

#### Quitline

Nair, U. S., Bell, M. L., Yuan, N. P., Wertheim, B. C., & Thomson, C. A. (2018). Associations between comorbid health conditions and quit outcomes among smokers enrolled in a state Quitline, Arizona, 2011-2016. *Public Health Reports*, *133*(2), 200-206. Available at: https://journals.sagepub.com/doi/full/10.1177/0033354918764903 *Compares a chronic health condition group (CHC) vs Mental health condition group (MHC), CHC and MHC group, vs no comorbid condition group*.

Vickerman, K. A., Schauer, G. L., Malarcher, A. M., Zhang, L., Mowery, P., & Nash, C. M. (2015). Quitline Use and Outcomes among Callers with and without Mental Health Conditions: A 7-Month Follow-Up Evaluation in Three States. BioMed Research International, 2015(1), 817298. Available at: https://onlinelibrary.wiley.com/doi/full/10.1155/2015/817298 *Compares users with and without a reported mental health condition*.

#### Psychological interventions

Heffner, J. L., Mull, K. E., Watson, N. L., McClure, J. B., & Bricker, J. B. (2018). Smokers with bipolar disorder, other affective disorders, and no mental health conditions: comparison of baseline characteristics and success at quitting in a large 12-month behavioural intervention randomized trial. *Drug and alcohol dependence*, *193*, 35-41. Available at: https://www.ncbi.nlm.nih.gov/pmc/articles/PMC6239897/ *Compares those with bipolar disorder vs other affective disorders vs no mental health conditions*.

Watson, N. L., Heffner, J. L., Mull, K. E., McClure, J. B., & Bricker, J. B. (2019). Comparing treatment acceptability and 12-month cessation rates in response to web-based smoking interventions among smokers who do and do not screen positive for affective disorders: secondary analysis. *Journal of Medical Internet Research*, *21*(6), e13500. Available at: https://www.jmir.org/2019/6/e13500/ *Compares a no affective disorder group vs six non-mutually exclusive subgroups: those with depression, posttraumatic stress disorder, panic disorder, generalized anxiety disorder, social anxiety disorder, and more than one affective disorder*.

Wen, S., Wiers, R. W., Boffo, M., Grasman, R. P., Pronk, T., & Larsen, H. (2021). Subtypes of smokers in a randomized controlled trial of a web-based smoking cessation program and their role in predicting intervention non-usage attrition: Implications for the development of tailored interventions. *Internet Interventions*, *26*, 100473. Available at: https://www.sciencedirect.com/science/article/pii/S2214782921001135 *Compares three clusters of smokers that differed on a broad range of characteristics – none diagnosed as depression and anxiety but relatively high scores on hopelessness, anxiety sensitivity, impulsivity, depression, and alcohol use*.

#### Pharmacological interventions

Wu, A. D., Gao, M., Aveyard, P., & Taylor, G. (2023). Smoking cessation and changes in anxiety and depression in adults with and without psychiatric disorders. *JAMA Network Open*, *6*(5), e2316111-e2316111. Available at: https://jamanetwork.com/journals/jamanetworkopen/article-abstract/2805442 *Compares people with and without psychiatric disorders*.

